# Mutations in EPG5 are associated with a wide spectrum of neurodevelopmental and neurodegenerative disorders

**DOI:** 10.1101/2024.06.12.24308722

**Authors:** Hormos Salimi Dafsari, Celine Deneubourg, Kritarth Singh, Reza Maroofian, Zita Suprenant, Ay Lin Kho, Neil J Ingham, Karen P Steel, Preethi Sheshadri, Franciska Baur, Lea Hentrich, Birgit Gerisch, Mina Zamani, Cesar Alvares, Ata Siddiqui, Haidar S Dafsari, Mehri Salari, Anthony Lang, Michael Harris, Alice Abdelaleem, Saeid Sadeghian, Reza Azizimalamiri, Hamid Galehdari, Gholamreza Shariati, Alireza Sedaghat, Jawaher Zeighami, Daniel Calame, Dana Marafi, Ruizhi Duan, Adrian Boehnke, Carrie Mohila, Dora Steel, Saurabh Chopra, Suvasini Sharma, Nicolai Kohlschmidt, Steffi Patzer, Afshin Saffari, Darius Ebrahimi-Fakhari, Büşra Eser Çavdartepe, Irene J Chang, Erika Beckman, Renate Peters, Andrew Paul Fennell, Bernice Lo, Luisa Averdunk, Felix Distelmaier, Martina Baethmann, Frances Elmslie, Kairit Joost, Sheela Nampoothiri, Dhanya Yesodharan, Hannah Mandel, Amy Kimball, Antonie D. Kline, Cyril Mignot, Boris Keren, Vincent Laugel, Katrin Õunap, Kalpana Devadathan, Frederique M.C. van Berkestijn, Arpana Silwal, Saskia Koene, Sumit Verma, Mohammed Yousuf Karim, Chahynez Boubidi, Majid Aziz, Gehad ElGhazali, Lauren Mattas, Mohammad Miryounesi, Farzad Hashemi-Gorji, Shahryar Alavi, Nayereh Nouri, Mehrdad Noruzinia, Saeedeh Kavousi, Arveen Kamath, Sandeep Jayawant, Russell Saneto, Nourelhoda A. Haridy, Pinar Ozkan Kart, Ali Cansu, Claire Beneteau, Kyra E. Stuurman, Martina Wilke, Tahsin Stefan Barakat, Homa Tajsharghi, Annarita Scardamaglia, Sadeq Vallian, Semra Hız, Ali Shoeibi, Reza Boostani, Narges Hashemi, Meisam Babaei, Norah Saleh Alsaleh, Julie Lander, Tania Attié-Bitach, Pauline Marzin, Dorota Wicher, Jessica I Gold, Mariana H G Monje, Dimitri Krainc, Niccolò Mencacci, Somayeh Bakhtiari, Michael Kruer, Emanuela Argilli, Elliott Sherr, Yalda Jamshidi, Ehsan Ghayoor Karimiani, Yiu Wing Sunny Cheung, Ivan Karin, Wendy K Chung, James R. Lupski, Manju A. Kurian, Jörg Dötsch, Jürgen-Christoph von Kleist-Retzow, Thomas Klopstock, Matias Wagner, Calvin Yip, Andreas Roos, Carlo Dionisi-Vici, Mathias Gautel, Michael R Duchen, Adam Antebi, Henry Houlden, Manolis Fanto, Heinz Jungbluth

**Affiliations:** Department of Pediatrics and Center for Rare Diseases, Faculty of Medicine and University Hospital Cologne, University of Cologne, Cologne, Germany; Max-Planck-Institute for Biology of Aging and Cologne Excellence Cluster for Ageing-associated Diseases, Cologne, Germany; Department of Paediatric Neurology, Evelina London Children’s Hospital, Guy’s & St Thomas’ NHS Foundation Trust, London, UK; Randall Centre for Cell and Molecular Biophysics, Muscle Signalling Section, Faculty of Life Sciences and Medicine (FoLSM), King’s College London, London, UK; Department of Basic and Clinical Neuroscience, IoPPN, King’s College London; UCL Consortium for Mitochondrial Research and Department of Cell and Developmental Biology, University College London, London, UK; Department of Neuromuscular Diseases, UCL Queen Square Institute of Neurology, London, UK; Vici Syndrome Foundation, Inc Silver Spring, Maryland, USA; Wolfson Sensory, Pain and Regeneration Centre, King’s College London, United Kingdom; Narges Medical Genetics and Prenatal Diagnosis Laboratory, Kianpars, Ahvaz, Iran; Department of Biology, Faculty of Science, Shahid Chamran University of Ahvaz, Ahvaz, Iran; Department of Radiology, Boston Children’s Hospital, 02115 Boston, Massachusetts, USA; Department of Radiology, Guy’s and Saint Thomas’ Hospitals NHS Trust, London, UK; Department of Neurology, Faculty of Medicine and University Hospital Cologne, University of Cologne, Cologne, Germany; Department of Neurology, Shahid Beheshti University of Medical Sciences Tehran Iran; Edmond J Safra Program in Parkinson’s Disease, Krembil Brain Institute, University Health Network and the Department of Medicine, University of Toronto, Canada; Departments of Neurology, Weill Cornell Medicine Qatar, Education City, P.O. 24144, Doha, Qatar; Medical Molecular Genetics, Institute Human Genetics and Genome Research, National Research Centre, Dokki, Egypt; Department of Pediatric Neurology, Golestan Medical, Educational, and Research Center, Ahvaz Jundishapur University of Medical Sciences, Ahvaz, Iran; Department of Medical Genetics, Faculty of Medicine, Ahvaz Jundishapur University of Medical Sciences, Ahvaz, Iran; Diabetes Research Center, Health Research Institute, Ahvaz Jundishapur University of Medical Sciences, Ahvaz, Iran; Departments of Pediatrics and Molecular and Human Genetics, Baylor College of Medicine, Houston, TX 77096 USA; Department of Pathology & Immunology, Baylor College of Medicine and Texas Children’s Hospital, Houston, TX, USA; Developmental Neurosciences, Zayed Centre for Research into Rare Disease in Children,UCL GOS-Institute of Child Health, London, UK; Indraprastha Apollo Hospital, New Delhi, India; Department of Pediatrics, Lady Hardinge Medical College and Associated Kalawati Saran Children’s Hospital, New Delhi, India; Institut fuer Klinische Genetik und Tumorgenetik Bonn, Bonn, Germany; Department of Pediatrics, Krankenhaus St. Elisabeth und St. Barbara, Halle (Saale), Germany; Division of Child Neurology and Inherited Metabolic Diseases, Heidelberg University Hospital, Heidelberg, Germany; Movement Disorders Program, Department of Neurology, Boston Children’s Hospital, Harvard Medical School, Boston, Massachusetts, USA; Department of Medical Genetics, Konya City Hospital, Konya, Turkey; Division of Medical Genetics, Department of Pediatrics, University of California San Francisco, San Francisco, California, USA; Division of Genetic Medicine, Seattle Children’s Hospital, Seattle, Washington, USA; Christliches Kinderhospital Osnabrück, Germany; Monash Genetics, Monash Health, Melbourne, Australia; Department of Paediatrics, Monash University, Melbourne, Australia; Research Branch, Sidra Medicine, Doha; College of Health and Life Sciences, Hamad Bin Khalifa University, Doha, Qatar; Department of General Pediatrics, Neonatology and Pediatric Cardiology, Medical Faculty, Heinrich-Heine-University, Düsseldorf, Germany; Department of Pediatrics, Hospital Dritter Orden, Munich, Germany; St George’s University Hospitals NHS foundation trust, London, UK; Faculty of Medicine at the University of Tartu, Tartu, Estland; Department of Pediatric Genetics, Amrita Institute of Medical Sciences & Research Centre, Cochin, India; Metabolic Disease Unit, Meyer Children’s Hospital, Haifa 31096, Israel.; Harvey Institute for Human Genetics, Greater Baltimore Medical Center, Baltimore, Maryland, USA; APHP, Hôpital Pitié-Salpêtrière, Département de Génétique, Centre de Reference Déficience Intellectuelle de Causes Rares, GRC UPMC Déficience Intellectuelle et Autisme, 75013 Paris, France; Service de Pédiatrie 1, Hôpital de Hautepierre, Hôpitaux Universitaires de Strasbourg, Strasbourg, France; Laboratoire de Génétique médicale, INSERM U1112, Institut de génétique médicale d’Alsace, Faculté de Médecine de Strasbourg, Hôpitaux Universitaires de Strasbourg, Strasbourg, France; Department of Clinical Genetics, Genetics and Personalized Medicine Clinic, Tartu University Hospital and Institute of Clinical Medicine, University of Tartu, Tartu, Estonia; Department of Paediatric Neurology, Government Medical College, Thiruvananthapuram, Kerala, India; Department of Pediatric Neurology, University Medical Center Utrecht, Utrecht, The Netherlands; St Bart’s Health NHS Trust, London, UK; Department of Clinical Genetics, Leiden University Medical Centre, Leiden, The Netherlands; Emory Children’s Hospital, Atlanta, Georgia, USA; Department of Pathology, Sidra Medicine, Doha; College of Medicine, Qatar University, Doha, Qatar; Department of Pediatrics A, Hussein Dey University Hospital Center, University of Algiers 1, Algiers, Algeria; Sheikh Khalifa Medical City, Abu Dhabi, UAE; Department of Medical Microbiology and Immunology, College of Medicine and Health Sciences, United Arab Emirates University, P.O. Box 15551, Al Ain, UAE; Stanford Children’s Hospital, Palo Alto, California, USA; Department of Medical Genetics, Faculty of Medicine, Shahid Beheshti University of Medical Sciences, Tehran, Iran; Genomic Research Center, Shahid Beheshti University of Medical Sciences, Tehran, Iran; Palindrome, Isfahan, Iran; Karyogen Lab, Isfahan, Iran; Department of Laboratory Medicine and Pathology, University of Washington, Seattle, Washington, USA; Department of Medical Genetics, Faculty of Medical Sciences, Tarbiat Modares University, Tehran, Iran; Cardiff and Vale UHB - AWMGS, Cardiff, UK; Oxford University Hospitals NHS Foundation Trust, Oxford, UK; Neuroscience Institute, Center for Integrated Brain Research, Department of Neurology/Division of Pediatric Neurology, Seattle Children’s Hospital, Seattle, WA, USA; Department of Neurology and Psychiatry, Faculty of Medicine, Assiut University, 71515, Assiut, Egypt; Karadeniz Technical University, Department of Pediatrics Neurology, Turkey; Genetics Department, Nantes University Hospital, Nantes, France; Deparment of Clinical Genetics, Erasmus MC University Medical Center, Rotterdam, The Netherlands; School of Health Science, Division Biomedicine and Translational Medicine, University of Skovde, SE-541 28 Skovde, Sweden; Department of Cell and Molecular Biology & Microbiology, Faculty of Science and Technology, University of Isfahan, Isfahan, Iran; Faculty of Medicine, Pediatric Neurology Department, Dokuz Eylül University, Izmir, Turkey; Department of Neurology, Mashhad University of Medical Sciences, Mashhad, Iran; Department of Pediatrics, School of Medicine, Mashhad University of Medical Sciences, Mashhad, Iran; Department of Pediatrics, North Khorasan University of Medical Sciences, Bojnurd, Iran; Department of Genetics and Precision Medicine, King Abdullah Specialized Children’s Hospital, King Abdullah International Medical Research Center, Ministry of National Guard Health Affairs, Riyadh, Saudi Arabia; Department of Pathology, University of Utah; Service de Médecine Génomique des Maladies Rares, Hôpital Necker-Enfants Malades, AP-HP, 75015 Paris, France; Department of Medical Genetics, Children’s Memorial Health Institute, Warsaw, Poland; Division of Medical Genetics, Department of Pediatrics, Cohen Children’s Medical Center, New Hyde Park, NY, USA; Department of Neurology, Northwestern University Feinberg School of Medicine, Chicago, Illinois 60611, USA; Pediatric Movement Disorders Program, Division of Pediatric Neurology, Barrow Neurological Institute, Phoenix Children’s Hospital, Phoenix, Arizona 85004, USA; Department of Neurology, University of California San Francisco Division of Hospital Medicine, San Francisco, California, USA; Molecular and Clinical Sciences Institute, St. George’s University of London, London, United Kingdom; Life Sciences Institute, Department of Biochemistry and Molecular Biology, The University of British Columbia, Vancouver, British Columbia, Canada; Friedrich-Baur-Institute, Department of Neurology, LMU University Hospital, Ludwig-Maximilians-Universität München, 80336 Munich, Germany; Department of Pediatrics, Boston Children’s Hospital and Harvard Medical School, Boston, MA 02115, USA; German Center for Neurodegenerative Diseases (DZNE), Munich, Germany; Munich Cluster for Systems Neurology (SyNergy), Munich, Germany; Institute of Human Genetics, School of Medicine and Health, Technische Universität München, Munich, Germany; Department of Pediatric Neurology, Centre for Neuromuscular Disorders, Centre for Translational Neuro- and Behavioral Sciences, University Duisburg-Essen, 45147, Essen, Germany; Department of Neurology, Medical Faculty and University Hospital Düsseldorf, Heinrich Heine University, 40225, Düsseldorf, Germany; Brain and Mind Research Institute, Children’s Hospital of Eastern Ontario Research Institute, Ottawa, ON, K1H 8L1, Canada; Division of Metabolic Diseases and Hepatology, Bambino Gesù Children’s Hospital, IRCCS, Piazza S. Onofrio 4, 00165, Rome, Italy

**Author notes:** **Correspondence to:** Heinz Jungbluth, MD, PhD, Department of Pediatric Neurology, Neuromuscular Service, Evelina’s Children Hospital, Guy’s & St. Thomas’ Hospital NHS Foundation Trust, London, United Kingdom. equal contribution (Hormos Salimi Dafsari, Celine Deneubourg, Kritarth Singh, Reza Maroofian). joint supervision (Michael R Duchen, Adam Antebi, Henry Houlden, Manolis Fanto, Heinz Jungbluth).

**Keywords:** Autophagy, *EPG5*, Neurodevelopmental disorders, Neurodegenerative disorders, Parkinson’s Disease

## Abstract

Autophagy is a fundamental and evolutionary conserved biological pathway with vital roles in intracellular quality control and homeostasis. The process of autophagy involves the engulfment of intracellular targets by autophagosomes and their delivery to the lysosome for digestion and recycling. We have previously reported recessive variants in *EPG5*, encoding for the ectopic P-granules 5 autophagy protein with a crucial role in autophagosome-lysosome fusion, as the cause of Vici syndrome (VS), a severe multisystem neurodevelopmental disorder defined by a combination of distinct clinical features including callosal agenesis, cataracts, cardiomyopathy, immunodeficiency, and hypopigmentation. Here, we present extensive novel genetic, clinical, neuroradiological and pathological features from the largest cohort of *EPG5*-related disorders reported to date, complemented by experimental findings from patient cells and models of EPG5 defects in *Caenorhabditis elegans* and *Mus musculus*. We identified 200 patients with recessive *EPG5* variants, 86 of them previously unpublished. The associated phenotypic spectrum ranged from antenatally lethal presentations and the classic VS phenotype (n=60) to much milder neurodevelopmental disorders with less specific manifestations (n=140). Myopathic features and epilepsy with variable progression were frequently observed. Novel manifestations included early-onset parkinsonism and dystonia with cognitive decline during adolescence, hereditary spastic paraplegia (HSPP), and myoclonus. Radiological findings included previously recognized *EPG5*-related features with callosal abnormalities and pontocerebellar hypoplasia, and a range of novel features suggesting an emerging continuum with disorders of brain iron accumulation or copper metabolism as well as HSPPs. Genotype-phenotype studies suggested a correlation between predicted residual EPG5 expression and clinical severity, especially regarding disease progression and survival. The Epg5 p.Gln331Arg knock-in mouse, a model of milder *EPG5*-related disorders, showed an age-related motor phenotype and impaired autophagic clearance in several brain regions mirroring those also affected in humans. In *Caenorhabditis elegans, epg-5* knockdown gave rise to neurodevelopmental features and motor impairment comparable to defects in parkinsonism-related genes, abnormal mitochondrial respiration, and impaired mitophagic clearance early in life. Cellular assays revealed impaired PINK1-Parkin dependent mitophagic clearance in patient fibroblasts. Our findings expand the phenotypic spectrum of *EPG5*-related disorders and indicate a life time continuum of disease that overlaps with other disorders of defective autophagy and intracellular trafficking. Our observations also suggest close links between early-onset neurodevelopmental and neurodegenerative conditions of later onset due to EPG5 defects, in particular dystonia and parkinsonism, highlighting the fundamental importance of dysfunctional autophagy in the pathophysiology of common neurodegenerative disorders.

## INTRODUCTION

Autophagy is a fundamental biological pathway conserved throughout evolution with important roles in intracellular quality control and homeostasis^1^. The process of its most common form of macroautophagy is characterized by the engulfment of intracellular targets by double-membraned structures, autophagosomes, and their delivery to the lysosome for digestion and recycling. Autophagy differs from the ubiquitin-proteasome system, another important intracellular housekeeping system, in its ability to process larger proteins and intracellular organelles such as mitochondria (termed ‘mitophagy’)^2^.

Following its original description in the 1960s, autophagy has been implicated non-specifically in a wide range of human disease, but monogenic primary autophagy defects in humans have only been recognized over the last decade^3^. Vici syndrome (VS), a multisystem disease defined by the key features of callosal agenesis, cataracts, hypopigmentation, cardiomyopathy and immunodeficiency, is the paradigmatic disorder of defective autophagy and associated with recessive variants in *EPG5*, encoding the ectopic P-granules 5 autophagy protein, a tethering factor with a key role in autophagosome-lysosome fusion^4-6^. Whilst more than 95% of individuals with the key features of VS carry detectable variants in *EPG5*, there is evidence that not all individuals with *EPG5* variants have classic VS, prompting the recently proposed term “*EPG5*-related disorders” (*EPG5*-RD)^7^. Most genotypes associated with classic VS have at least one truncating *EPG5* variant, whereas the few reported milder cases carried at least one *EPG5* missense variant at compound heterozygous or homozygous state^7,8^. The most common recurrent variant is the p.Gln336Arg founder variant prevalent in Ashkenazi populations^9,10^ that was originally identified in homozygosity in a patient with learning difficulties, epilepsy and a myopathy but no other features of classic VS^11^.

Moreover, preliminary observation in families of patients with *EPG5*-RDs suggest an increased risk for subtle VS manifestations (in particular cataracts and focal hypopigmentation), certain cancers, and movement disorders including Parkinson’s disease (PD) in (putative) heterozygous *EPG5* variant carriers^11^. These findings may indicate a gene dosage effect of EPG5 function, similarly to observations in *GBA*-related Gaucher disease where heterozygous carriers may develop early-onset parkinsonism^12^. Along similar lines, emerging evidence from other autophagy and intracellular trafficking disorders suggests that these conditions reflect a lifetime disease continuum depending on the degree of protein dysfunction^13^.

In this study we report the expanding phenotypic spectrum associated with *EPG5* variants identified through either phenotype-driven (i.e. targeted *EPG5* testing prompted by classic VS features) or reverse-phenotyping (i.e. exome/genome sequencing in non-specific neurodevelopmental disorders) approaches. Clinical and complementary experimental data from mouse, worm and cellular models suggest a spectrum of neurodevelopmental and neurodegenerative phenotypes linked to EPG5 defects.

## MATERIALS AND METHODS

Key methods for the different parts of our study are outlined below and additional information is detailed in Supplementary File 1.

### Clinical investigations in patients with *EPG5*-RDs

Patients with biallelic *EPG5* variants were recruited from multi-center studies via established collaborations, web platforms (GeneMatcher, Varsome), and national studies on rare pediatric neurological diseases in the European Union (ERN ITHACA), Germany (ESNEK), and the United Kingdom (100K Genomes Project, DDD study)^14-17^. The diagnosis of classic VS was based on the presence of at least 5 out of 7 features, as defined by Dionisi-Vici and subsequently refined by Byrne et al. (2016)^5,6^.

Deep phenotyping was performed based on patient histories, muscle biopsies, and brain magnetic resonance imaging (MRI) studies obtained as part of the routine diagnostic process and patient care. Brain MRIs were assessed independently by two expert pediatric radiologists (CA, AS). Video captured movement disorder phenotypes were assessed independently by three movement disorders experts (AL, MS, HD), converging independently on an expert opinion. Patient data was collected using a standardized proforma including 207 items (Supplementary File 2). In addition, we reviewed the literature for all previously published patients with pathogenic *EPG5* variants and contacted lead authors for up-to-date information.

The study was approved by the ethics committee of the Medical Faculty, University of Cologne (20-1711) and other regional institutional review boards^18^. All patients and/or their legal guardians gave informed consent to anonymized publication and use of recognizable photographs/videos where applicable.

### Genomic investigations in patients with *EPG5*-RDs

*EPG5* variant screening was performed by targeted single-gene testing in patients with diagnostic features of VS, or by exome/genome sequencing as previously published^19^. Dideoxy sequencing was performed to test for co-segregation of variants with the disorder in individual families.

*EPG5* variants identified were included if classified as pathogenic or likely pathogenic classifications according to ACMG criteria. Variants of unclear significance were additionally subjected to a rigorous additional analysis using various bioinformatic pathogenicity prediction tools (CADD, MutPred, spliceAI, other score values), taking into account reports in the gnomAD healthy population database^20-22^.

### Generation and characterization of an *Epg5*^*Q331R*^ knock-in mouse model

*Epg5*^*Q331R*^knock-in mice were generated by Taconic through CRISPR/Cas9-mediated gene editing in a C57BL/6N background (C57BL/6NTac^-^*Epg5*^*em4827(Q331R)Tac*^). The colony was maintained on a 12h light/dark cycle and had unlimited access to food and water (PicoLab Rodent Diet 20 (5053, LabDiet). Cage enrichment consisted of cardboard houses, tubes, and nesting materials. All procedures were carried out under the Animals (Scientific Procedures) Act of 1986 (United Kingdom) under appropriate Home Office project licences.

To characterize Epg5 levels and the autophagy defect in mice, the murine brain was divided in four separate regions (forebrain, midbrain, cerebellum, and brainstem) from which RNA and protein samples were extracted and western blot analysis or RT-qPCR was performed. Standardized assessments were used to characterize the behavioural phenotype of the mice at 1.5 months (6±1 weeks), defined as “early stage”, and 11±1 months, defined as “endstage”. In line with common VS manifestations, we also assessed potential effects of Epg5 defects on hearing and muscle. Supplementary File 1 details methods on behavioural analyses.

### Generation and characterization of a *C. elegans* epg-5 knockdown model

Nematodes were cultured at 20°C on nematode growth medium (NGM) agar plates using standard techniques. For RNAi experiments, worms were grown on *Escherichia coli* HT115 strain transformed with L4440 vector for expression of double-stranded RNA (dsRNA) obtained from the Ahringer library^23^. RNAi bacteria were grown overnight at 37°C in Luria Broth with 50□μg mL^−1^ ampicillin and 10□μg mL^−1^ tetracycline. The cultures were spun down at 4,000□r.p.m. at 4°C for 10□min. 500□μL of one-fold concentrated culture was seeded onto NGM plates containing 1M isopropyl β-d-1-thiogalactopyranoside (IPTG) to induce dsRNA expression. All RNAi feeding was carried out from egg-on. To investigate for motor phenotypes, we carried out locomotion analyses^24^ in worms with knockdown in *epg-5/EPG5*, the autophagosome-lysosome maturation gene *rab-7/RAB7* as a positive control for stalled late autophagy, the vacuolar fusion gene *ccz-1/CCZ1* as a positive control for stalled fusion, and the mitophagy gene *pdr-1/PRKN* as a positive control for a PD-related disease gene.

To assess mitophagy flux, we used the *C. elegans* strain IR621:N2;*Ex002*[p_*lgg-1*_DsRed::LGG-1] for measurement of the autophagosome marker LGG-1/LC3 and mitophagy marker DCT-1, the ortholog of mammalian BNIP3 and BNIP3L/NIX^25,26^. Worms were grown on RNAi bacteria from egg-on. Three independent biological replicates of 10 hand-picked adult day 1 nematodes each were used for microscopy. Images of nematodes with control RNAi (*luci*) and *epg-5i* were taken with Andor Dragonfly at 60x magnification at day 1 adulthood. Punctae counting was performed manually while analysing researchers were blinded to image titles.

Supplementary File 1 details methods concerning neuronal morphology, locomotion analyses and oxygen consumption rate measurements.

### Cellular autophagy and mitophagy studies in patient fibroblasts and Epg5^Q331R^ mouse embryonic fibroblasts (MEFs)

Fibroblasts carrying p.Gln336Arg and p.Arg1621Gln homozygous EPG5 mutation were isolated from skin biopsies obtained from patients as part of the diagnostic process. Healthy control fibroblasts were obtained from the MRC Centre for Neuromuscular Disorders Biobank, London. MEFs were isolated from E15 embryos and immortalized using transformation with the SV40 plasmid (pBABE-puro SV40 LT, Addgene, #13970). Fibroblasts and MEFs were cultured and maintained in Dulbecco’s modified Eagle’s medium (Gibco #10566016) supplemented with 10% foetal bovine serum (Gibco #16140071), and 1% Antibiotic-Antimycotic (Gibco #15240096) and incubated at 37°C with 5% CO_2_. All cell cultures were routinely checked for mycoplasma using MycoAlert™ Mycoplasma Detection Kit (Lonza #LT07-118).

Fibroblasts were either transduced with mito-Keima (Addgene plasmid #56018) lentivirus or transfected with GFP-Parkin (Addgene plasmid #45875) and mRFP-GFP-LC3 (Addgene plasmid #21074) using Human Dermal Fibroblasts Nucleofector Kit (Lonza #VPD-1001) according to the manufacturer’s instructions. Cells were treated as indicated and imaged on LSM 880 Airyscan microscope using plan Apochromat 63x/1.4 oil DIC objective lens at 37°C. At least 8 z-stacks with 0.45μm thickness were acquired using Zen Black software (Carl Zeiss). High (543/458) ratio areas and total mitochondrial area were binarized, segmented and quantified in Fiji and used as a mitophagy index. LC3 and GFP-Parkin images were binarized and puncta/cell was quantified in Fiji.

## RESULTS

### Clinical findings

We obtained data of 200 patients with pathogenic *EPG5* variants from 139 families. In addition to 86 novel cases, we reviewed 114 published cases and contacted lead authors to obtain follow-up data on 88/114 patients.

There was a known history of consanguinity in 96 cases from 53 families. We found positive family histories with (putative) *EPG5* variant carriers positive for cancer (n=29, mostly breast and gastric cancer, melanoma, leukemia/lymphoma), neurological disorders including Parkinson’s disease (n=25), twin pregnancies (n=10), and vitiligo (n=7).

Amongst the key clinical diagnostic criteria for classic VS, callosal agenesis was most common (see below) but other features were more variable^6,11^. The most common non-neurological findings included failure-to-thrive (n=121) and hypopigmentation (n=107) relative to the familial and/or ethnic background. In addition to cataracts (n=78), optic atrophy (n=50) was another common ophthalmological feature. Sensorineural hearing loss, another feature in VS^27^, was found in 39 cases. Cardiac involvement included mostly dilated cardiomyopathy (n=72) but also a range of congenital heart defects (n=22). Microcephaly was observed in 106 cases (congenital in 18 cases and acquired in the remainder) but was less common in patients at the milder end of the spectrum. Microcephaly was often associated with other dysmorphic features (n=79) such as micro-/retrognathia, small bitemporal diameter, prominent upper lip, high-arched palate, low-set and posteriorly rotated ears, frontal bossing, and arachnodactyly. 38 patients had an established immunodeficiency confirmed in laboratory investigations, although a disproportionately large number of opportunistic infections and cardiorespiratory failure associated with febrile illness in patients without formal immunologic diagnostics suggested that primary immunodeficiency may be more common in *EPG5*-RDs than formally recognized. Pulmonary involvement was one of the most common manifestations present in 90 patients and included recurrent respiratory infections, aspirations and, less frequently, aspirations and apnoea. Hepatic involvement was found in 70 cases, including hepatosplenomegaly and elevated transaminase levels.

#### Neurological features

Delay of motor, speech, and/or cognitive milestones were reported in all patients. Termination of pregnancy or death in early infancy prevented the assessment of motor development in nine cases. Significant motor developmental delay was found in 183 cases surviving beyond early infancy. Epilepsy was the most common neurological feature, present in 104 and associated with a clearly progressive course in 28 patients. Five patients were reported with generalized motor status epilepticus and non-convulsive status. A detailed analysis of *EPG5*-related epilepsy has been reported elsewhere^28^.

A substantial proportion presented with a progressive neurodegenerative disorder with a strong movement component on the background of a non-specific neurodevelopmental disorder. In particular, movement disorders were found in 64 patients without primary clinical suspicion of VS, featuring spasticity (n=46, of which n=29 had isolated/pure spastic paraplegia), dystonia and/or parkinsonism (n=20), myoclonus (n=18), or a combination of the above. In most of these patients, genetic testing (mainly through exome/genome sequencing) was only requested after the onset of the movement disorder, leading to significant diagnostic delays. Ten cases showed mild neurodevelopmental delay in early childhood but developed rapidly progressive ‘atypical parkinsonism’ of sudden onset during adolescence, with additional symptoms not typically associated with sporadic parkinsonism, including preceding dystonia and spasticity, and rapid cognitive decline. Box 1 provides clinical vignettes and Figure 1 illustrates relevant brain imaging findings from illustrative cases within this subgroup. In addition, two particularly severely affected siblings compound heterozygous for *EPG5* variants (c.5869+1G>A;p.Trp1989Ter) presented with myoclonus from birth, followed by dystonia from four weeks, early-onset parkinsonism at three months, progressive infantile epileptic encephalopathy, and demise at around toddler age.

**Figure 1.**
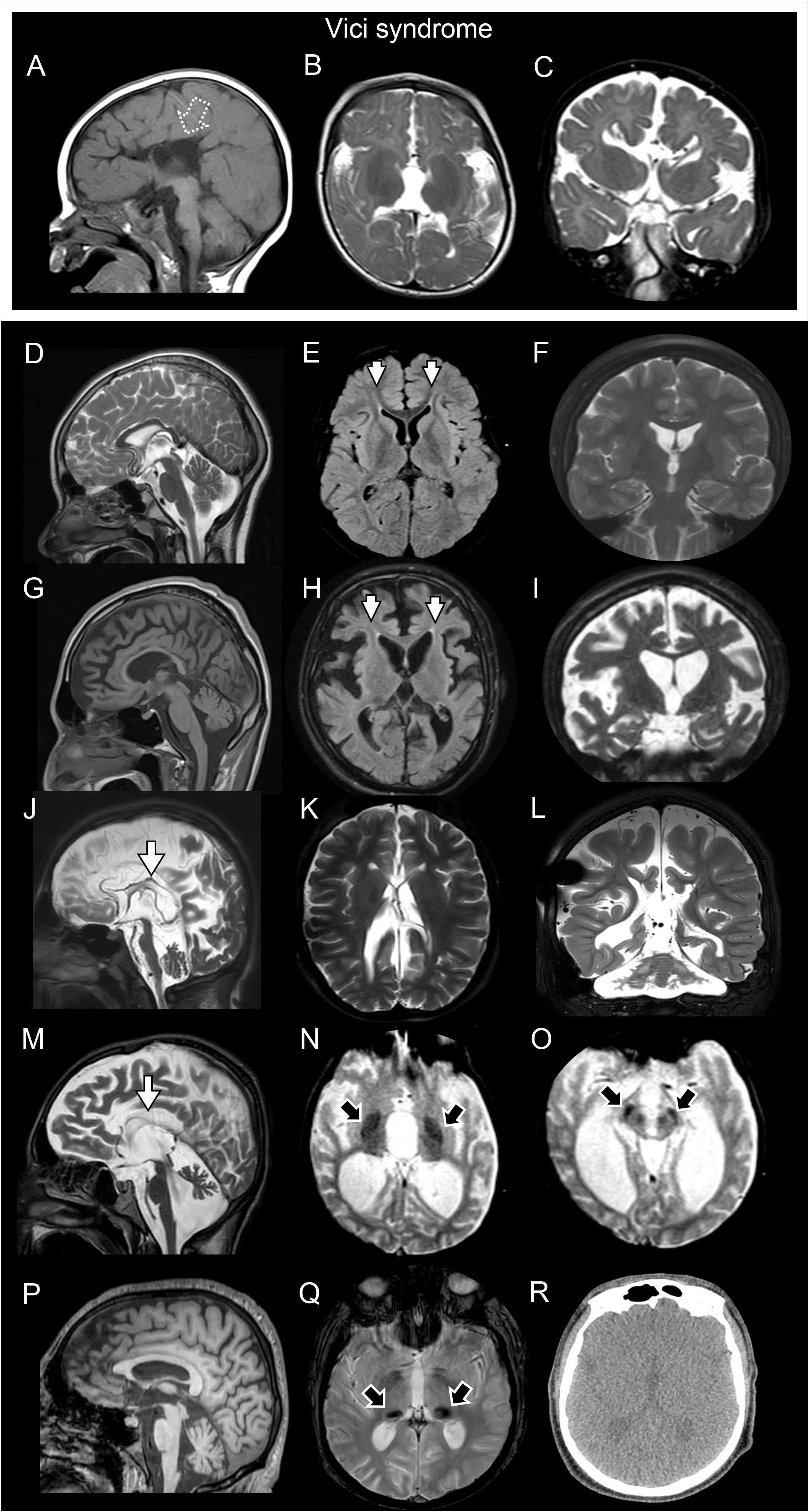
Neuroradiological spectrum in patients with EPG5-related disorders. Brain magnet resonance imaging of 6 different patients with *EPG5*-related disorders including 1 patient (A-C) with Vici Syndrome and 5 patients (D-R) with atypical presentations. Different degree of degenerative involvement of the brain varying from mild forms where there is selective involvement of the fornix minors (short arrows, E and H), without or with mild corpus callosum atrophy and atrophy of the brain to more severe forms where there is extremely thin and partially visualized corpus callosum (arrows, J-L) and cerebellar atrophy. Additional features including iron / micromineral deposition were noted in the globus pallidi (black arrow, N), and substantia nigra and red nuclei (black arrow, O) and also as an isolated feature more prominent in the pulvinar of the thalami (black arrows, Q).

#### Neuroradiological findings

We obtained information from a total of 174 MRI and/or CT scans from patients with *EPG5*-RDs. Brain MRI features in this cohort were suggestive of a structural neurodevelopmental disorder with abnormalities of the corpus callosum (n=169, including predominantly callosal agenesis but also rarer cases of callosal dysgenesis), optic atrophy (n=48), and (ponto-)cerebellar hypoplasia (n=46). Less frequent features included thalamic involvement (n=14), schizencephaly (n=4), and heterotopias (n=3). Other notable features included the ‘ear of the Lynx sign’, a FLAIR cone shaped periventricular hyperintensity usually associated with spastic paraplegia (n=3), and iron accumulation in basal ganglia (n=3) (Figure 1N-O,Q).

#### Neuromuscular features

Mostly generalized muscle weakness suggestive of a myopathy was observed in 131 cases. CK levels when measured were variable but typically elevated, ranging from 50 to 2130 IU/L. Muscle biopsies were performed in 39 cases and showed a wide range of features, including increased fibre size variability, type 1 predominance, increased vacuolization and lipid accumulation as the most common findings. Muscle biopsies in 12 patients indicated fibre size variation and/or cores. A range of mitochondrial abnormalities was prominent on electron microscopy, including abnormalities of mitochondrial number, size, distribution and internal structure, including abnormal cristae and inclusions.

### Molecular genetic findings

We obtained molecular genetic data from 200 cases, of which 121 cases carried homozygous variants and 79 cases compound heterozygous *EPG5* variants. In total, 110 cases carried truncating and/or splice site variants on both alleles (‘bi-truncating’), 33 cases had a combination of one truncating/splice site variant and one missense variant (‘mixed’), and 57 cases had missense variants on both alleles (‘bi-missense’). Type and distribution of *EPG5* variants identified and protein models are illustrated in Figure 2 and Supplementary File 1. Results of *in silico* variant assessments are summarized in Supplementary File 3.

**Figure 2.**
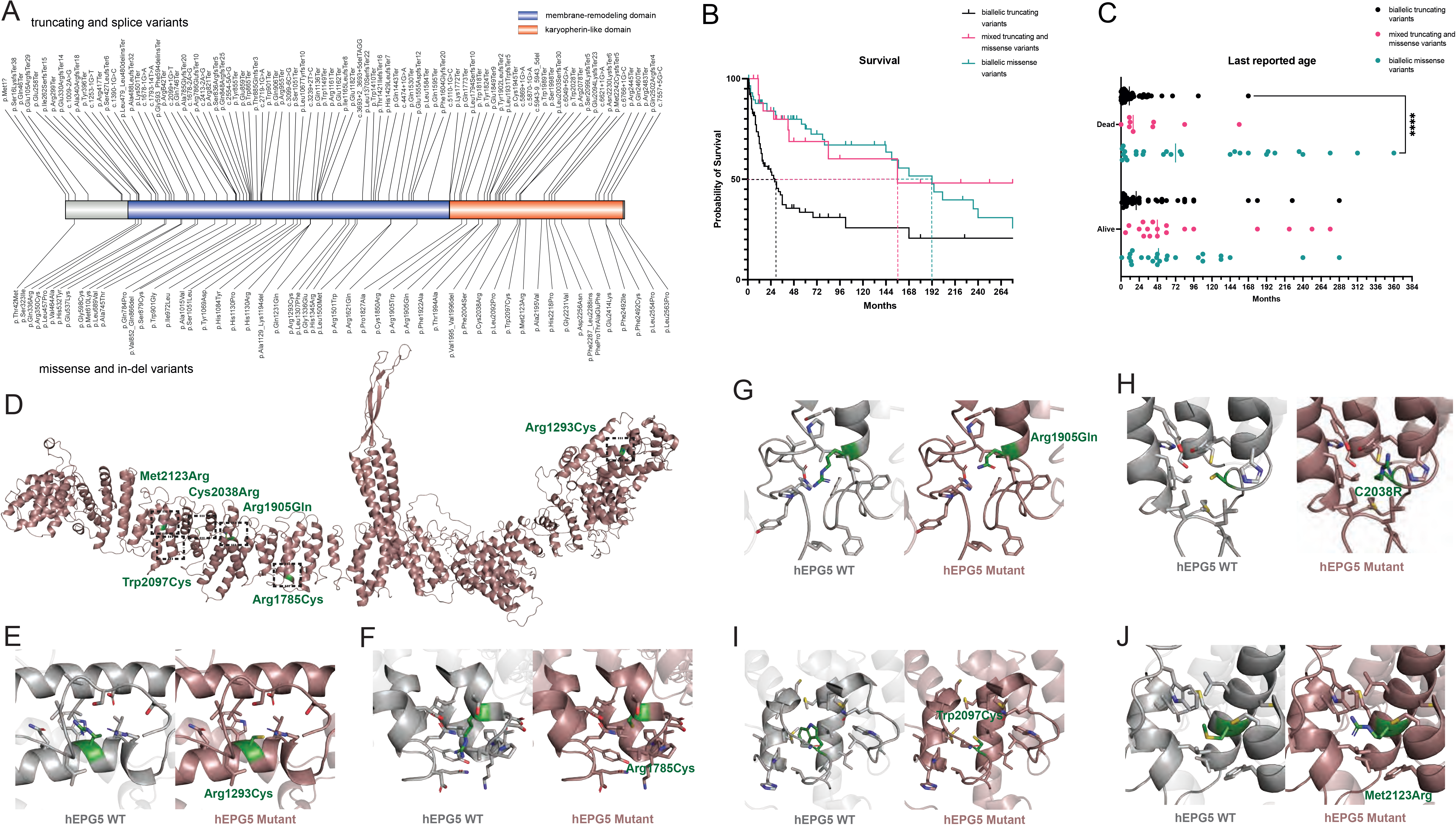
*EPG5* mutational spectrum and genotype-phenotype correlations in our cohort. (A) Protein structure with *EPG5* variants from our cohort of 200 patients. Truncating and splice site variants are shown above the schematic representation of the protein, missense and in-del variants are picture below (image created with IBS 2.0. (B) Kaplan-Meier survival curve of all patients in the three subgroups: biallelic truncating (and splice site) variants, mixed truncating and missense variants, and biallelic missense variants. Median life expectancy noted for each curve. (D) Last reported age for each of the groups: biallelic truncating (and splice site) variants, mixed truncating and missense variants, and biallelic missense variants. Overview (E) and specific effects (F-I) of *EPG5* missense variants in relation to the EPG5 protein model, indicating changes in protein structure. More detailed descriptions in Supplementary File 1.

Based on the classification of genotypes into three subgroups as outlined above, we could also establish clear genotype-phenotype correlations: Where the initial diagnosis was classic VS (n=60), *EPG5* variants were bi-truncating in 55%, mixed in 16.5% and bi-missense in 28.5%. Where the initial diagnosis was a non-specific neurodevelopmental disorder (n=140), *EPG5* variants were bi-truncating in 22%, mixed in 13% and bi-missense in 65% of patients. Analysis of variance (ANOVA) confirmed a significant enhancement of bi-truncating variants in classic VS patients.

Median life expectancy showed also significant differences (p<0.001, Log-rank Mantel-Cox test) between groups defined by genotype (Figure 2B-C). In particular, median life expectancy was 28 months in patients with bi-truncating *EPG5* variants, 156 months in patients with mixed truncating and missense *EPG5* variants and 192 months in patients with *EPG5* bi-missense variants. Within the first month of life, 42% of patients with bi-truncating variant are at risk of death, compared to only 15% of patients with bi-missense variants.

Eight patients were homozygous for the *EPG5* p.Gln336Arg variant, a likely Ashkenazi founder variant^9^ and had a relatively milder/attenuated phenotype, whereas those compound heterozygous for p.Gln336Arg and another more deleterious *EPG5* variant (n=3) had a more severe disorder intermediate between classic VS and milder phenotypes.

### An *Epg5*^*Q331R*^ knock-in mouse model displays a mild, age-dependent motor phenotype

We generated a new p.Gln331Arg/Q331R knock-in mouse model, corresponding to the recurring *EPG5*:p.Gln336Arg mutation associated with an attenuated phenotype^9^. Amino acid residue 331 is encoded by the last codon of exon 2, and we investigated whether the mutation resulted in aberrant *Epg5* mRNA splicing. The nucleotide change caused retention in intron 2, resulting in two different isoforms, one containing an extra 13 nucleotides of intron 2 and the other containing 386 nucleotides (Figure 3A). Both isoforms resulted in frameshifts that led to the introduction of stop codons and were predicted to be degraded. Importantly, correctly spliced mRNA that just contains the point mutation was also detected at lower levels (Figure 3A, Supplementary File 4A). Investigating *Epg5* mRNA levels by qPCR showed a significant mRNA reduction in midbrain and brainstem, with a similar but not significant trend in other brain areas (Figure 3B). Low mRNA levels were still present, likely due to the presence of normally spliced mRNA carrying only the point mutation.

**Figure 3.**
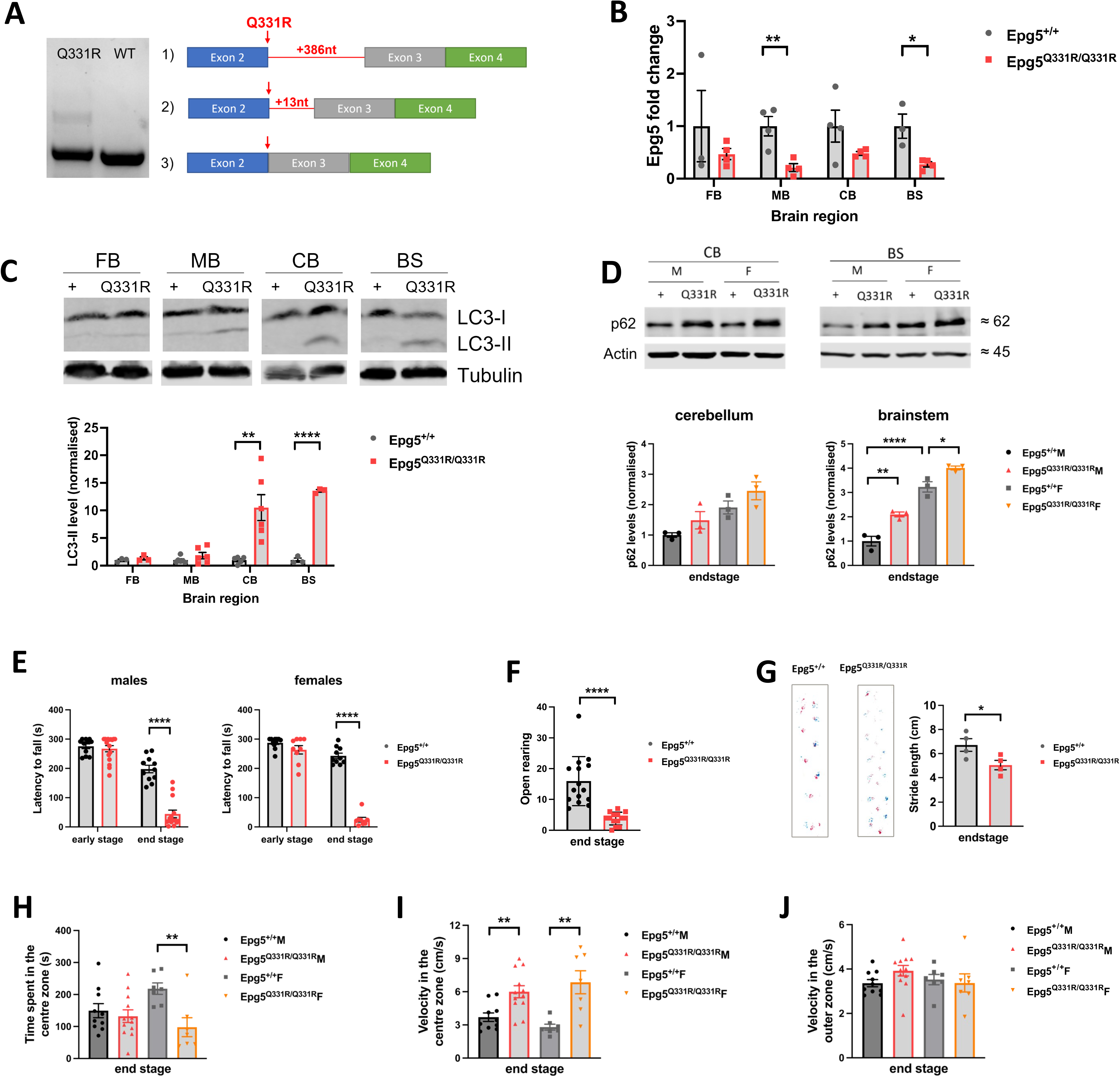
Key features from the *Epg5*^*Q331R*^ knock-in mouse model. (A) The murine *Epg5*^*Q331R*^ mutation (corresponding to the human *EPG5* p.Gln336Arg variant) causes splicing defects that result in two incorrectly spliced Epg5 isoforms. Normally spliced *Epg5* containing the point mutation is also present. (B) *Epg5* mRNA levels are significantly reduced in the midbrain and brainstem of *Epg5*^*Q331R*^ KI mice. (C) A strong increase in LC3-II is seen in the cerebellum and brainstem of the mutant mice. (D) A mild increase in p62 is seen in the cerebellum and a strong increase is seen in the brainstem of the mutant mice. (E) *Epg5*^*Q331R*^ KI mice show no motor phenotype at an early stage but fall off the rotating rod much faster than wildtype mice at endstage (∼12 months). (F) At endstage, mutant mice also show reduced rearing behaviour in an open arena compared to wildtype mice. (G) Stride length is significantly shortened in KI mice at endstage on RotaRod analysis. (H) Female *Epg5*^*Q331R*^ KI mice spent significantly less time in the centre zone of an arena than wildtype mice, while there is no significant difference between males. (I) *Epg5*^*Q331R*^ KI mice move through the virtually indicated centre zone of an arena at higher speed than wildtype mice, while (J) *Epg5*^*Q331R*^ KI mice do not move faster through the outer zone, indicating an anxiety phenotype.

The autophagosome marker LC3-II was strongly increased in the cerebellum and brainstem, corresponding to brain areas also preferentially affected in humans, but not in the forebrain and the midbrain (Figure 3C). p62 levels also appeared to be increased in those brain regions (Figure 3D), with substantially higher levels in females compared to males.

*Epg5*^*Q331R*^ homozygous mice appeared to develop normally and maintained a normal weight throughout early life (Supplementary File 4B). The mutant mice did not show any motor phenotype on a rotarod at 6 weeks of age (Figure 3E). At ∼12 months of age (endstage), however, a motor-related phenotype developed with clear deficits in latency to fall, open rearing behaviour, and stride length (Figure 3E-G). A grip strength assay was performed to investigate whether this was due to a loss of muscle strength as observed in the *Epg5* KO mice^29^, however, this was not confirmed in the *Epg5*^*Q331R*^ homozygous mice (Supplementary File 4C), suggesting that the motor phenotype observed in the *Epg5*^*Q331R*^ mice is likely due to reduced balance control and/or coordination rather than reduced neuromuscular strength. Typical features of vestibulocochlear dysfunction such as head bobbing, curling behaviour and hearing impairment (raised thresholds for otoacoustic emissions or auditory brainstem responses, Supplementary File 4D-G) were not observed, suggesting that the balance and/or coordination issues with impaired Preyer reflex and contact righting time may rather be a consequence of the observed cerebellar and brainstem alterations rather than of primary vestibular dysfunction (Supplementary File 4H-I).

In addition, *Epg5*^*Q331R*^ mice showed a mild increase in anxiety at endstage. In particular, mutant females spent significantly less time in the centre zone of an open arena compared to wildtype mice, suggesting stronger avoidance of perceived danger (Figure 3H). *Epg5*^*Q331R*^ mice also appeared to move through the centre zone at increased speed compared to wildtype but not in the outer zone of the arena (Figure 3I-J), again suggesting higher anxiety levels.

In conclusion the *Epg5*^*Q331R*^ homozygous mice display an age-related motor phenotype different from that previously reported in an *Epg5* ko mouse model, consistent with the expanded and milder spectrum of *EPG5*-RDs also observed in humans with the same genetic alteration.

### *epg-5* knockdown in *C. elegans* causes impaired mitophagy

To explore neurodevelopmental abnormalities associated with EPG5 deficiency further *in vivo*, we resorted to simpler model organisms, however, a full knockout of *epg5* in *Drosophila melanogaster* failed to reveal any neurodevelopmental defects^28^. Next, we conducted knockdown of *EPG5/epg-5* in a *Caenorhabditis elegans* model and observed aberrant neuronal morphology in quaternary dendrites in a reporter strain with *epg-5* egg-on knockdown for somatosensory neurons as early as larval stage 3 (Figure 4A).

**Figure 4.**
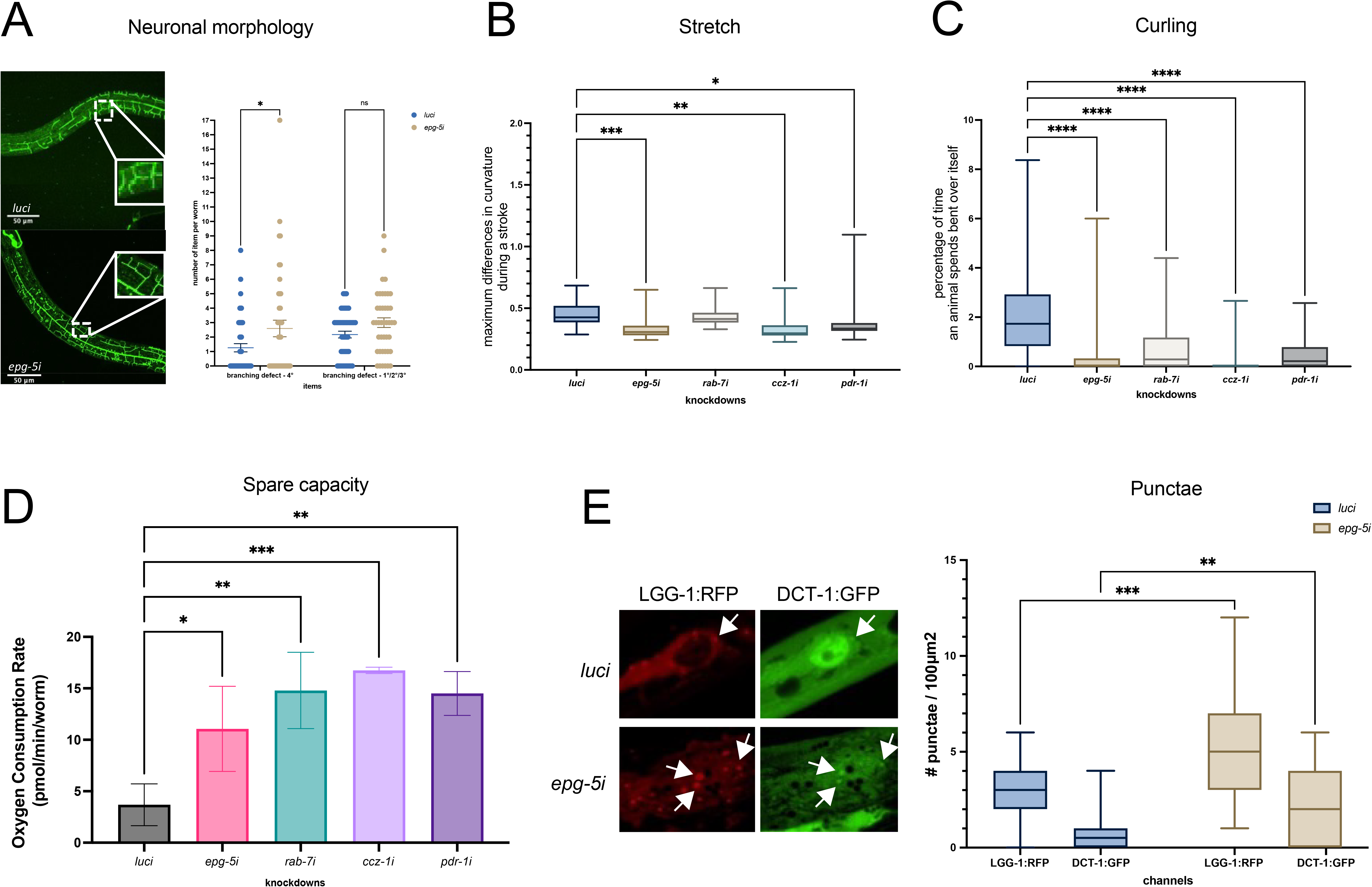
*C. elegans* model of EPG5 dysfunction. Neuronal morphology, motor phenotype, mitochondrial oxygen consumption, and mitophagy flux in *Caenorhabditis elegans* knockdown of *epg-5*. (A) Aberrant neuronal morphology in quartenary dendrites during larval stage 3 indicates a neurodevelopmental defect for *epg-5* knockdown worms. (B) Analysis of stretch showed significantly decreased maximum differences in curvature per stroke in *epg-5i, ccz-1i*, and *pdr-1i*, indicating flatter body bends, demonstrated by pooled data from three biological replicates of n = 10 worms. (C) Analysis of curling showed significantly decreased percentage of time spent in bent-over shapes in *epg-5i, rab-7i, ccz-1i*, and *pdr-1i* worms, indicating much flatter movements, demonstrated by pooled data from three biological replicates of n = 10 worms. (D) Measurement of oxygen consumption rate by Seahorse Respirometer revealed an increase in spare capacity in *epg-5i, rab-7i, ccz-1i*, and *pdr-1i*, indicating an excess of uncoupled capacity of the respiratory electron transport chain not being used in basal respiration. (E) Microscopy of mitophagy flux revealed an increase in LGG-1:RFP and DCT-1:GFP puncta in body wall muscle cells when compared to *luci* control. Quantification of punctae per 100 μm^2^ after counting by eye, shown as pooled data from three biological replicates of n = 10 worms. Statistical significance levels: * p ≤ 0.05, ** p ≤ 0.01, *** p ≤ 0.001 as determined by ANOVA.

Prompted by our observations of parkinsonian phenotypes in patients, we investigated the *in vivo* organismic consequences of *epg-5* knockdown in *C. elegans* taking into account the experience from existing *C. elegans* models of Parkinson Disease (PD). Defects in PD genes have previously been shown to result in decreased body bends and lower thrashing in *C. elegans*^30^. Knockdown of autophagosome-lysosome fusion genes (*epg-5, rab-7, ccz-1*) led to reduced locomotion similar to *pdr-1/PRKN* knockdown (Figure 4B-C), suggesting a hypokinetic phenotype also for the *epg-5* knockdown.

Given the link between PD and mitophagy regulation, we performed an oxygen consumption assay in worms to investigate for mitochondrial function abnormalities associated with *epg5*-deficiency. Knockdown of *pdr-1/PRKN* in *epg-5* wildtype resulted in an increase in the differential between maximum and basal oxygen consumption rate (‘spare capacity’), when compared to control *luci* worms, indicating an excess of uncoupled capacity of the respiratory electron transport chain not being used in basal respiration. We observed that knockdown of autophagosome-lysosome fusion genes *epg-5* and *ccz-1* in worms also led to significant increases in spare capacity when compared to controls (Figure 4D). Finally, to investigate for stalled mitophagy flux, we examined *epg-5* knockdown in a mitophagy reporter strain that indicated increased punctae of LGG-1:RFP and DCT-1:GFP in body wall muscles from *epg-5i* worms when compared to control (Figure 4E).

In conclusion, *epg-5* RNAi in *C. elegans* is associated with neurodevelopmental defects, motor defects, and mitochondrial and mitophagy alterations similar to those caused by abnormalities of PD genes in the same model organism.

### Cellular *EPG5* models show impaired PINK/Parkin-dependent mitophagy

Based on histopathological evidence for a mitochondrial cytopathy as well as stalled mitophagy flux in *epg-5* knockdown worms, we investigated for mitochondrial abnormalities in fibroblasts from patients with milder *EPG5*-RDs, homozygous for p.Gln336Arg and p.Arg1621Gln, respectively, both previously reported^9,11^.

p62/SQSTM1, NDP52 and LC3-II immunoblotting showed higher levels of these proteins in patient fibroblasts under basal condition (Figure 5A) that increased even further following treatment with rapamycin (prompting autophagy flux) and bafilomycin (blocking autophagy flux) (Figure 5B and Supplementary File 5A-B), confirming the reported role of EPG5 in autophagosome-lysosome fusion^4^. We further analysed autophagosome-lysosome fusion in EPG5-defective cells transfected with tandem-fluorescent mRFP-GFP-LC3 (Supplementary File 5C) and demonstrated an almost complete merge of GFP and RFP signals under all conditions indicating autophagosome accumulation before fusion (Supplementary File 5D).

**Figure 5.**
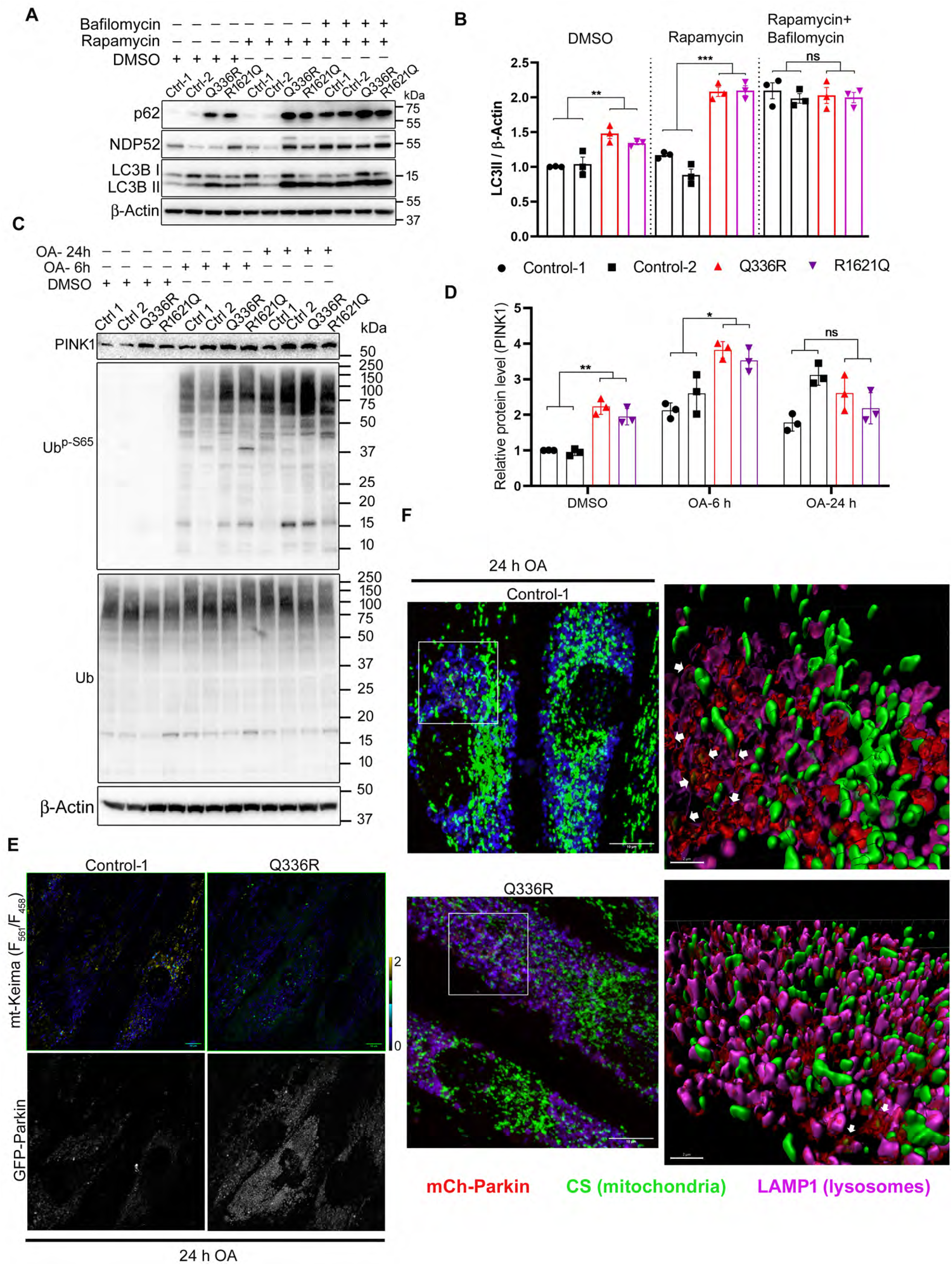
Impaired autophagic flux and mitophagy defects in *EPG5*-mutated patient cells and MEFs. (A) Immunoblot showing accumulation of p62, NDP52 and LC3II in control and patient fibroblasts after 24 h of treatment with rapamycin (100 nM) and/or bafilomycin (200 nM). (B) Quantification of LC3II/β-Actin ratio in A. (C) Immunoblot showing accumulation of PINK1 levels and its activity (phospho-Ub^S65^) in both Q336R and R1621Q fibroblasts in response to oligomycin (2.5 μM) and antimycin (2.0 μM) (OA) treatment for indicated time. Ub blot shows the total ubiquitin levels. (D) Quantification of the PINK1 levels in C. (E) Representative images of fibroblast cells co-transfected with mito-Keima (ratiometric) and GFP-Parkin (grey scale) after treatment with OA for 24 h. Scale bars, 10 μm. (F) Representative immunofluorescence images of mCherry-Parkin transfected fibroblasts immunostained for citrate synthase (CS) and LAMP1. 3D reconstruction of inset in Control-1 shows Parkin-CS puncta inside the LAMP1-positive vesicles (white arrows). These structures were counted as ‘mitophagy events’ which was further plotted. Scale bar: overview, 10 μm; inset, 2 μm. Data in B) and D) are represented as mean ± s.d. from three independent experiments. *P < 0.05, **P < 0.005, ***P < 0.001, ****P < 0.0001 (two-way ANOVA). ns: not significant.

As accumulation of p62 and the redox sensor NDP52 indicated a role of EPG5 in mitophagy-mediated mitochondrial quality control, we investigated EPG5 during mitophagy induction and observed co-localization of EPG5 and fragmented mitochondria in response to mitochondrial depolarization induced by oligomycin and antimycin-induced mitochondrial depolarization (Supplementary File 5E). To delineate the mechanism of mitophagy inhibition further, we analysed the accumulation of the mitochondrial kinase PINK1 following mitochondrial depolarization by immunoblotting. Interestingly, PINK1 levels were already significantly elevated in p.Gln336Arg and p.Arg1621Gln cells at baseline and increased even further after 6h treatment with oligomycin and antimycin (‘OA’, indicative of early mitophagy, Figure 5C-D). Levels of ubiquitin phosphorylation at Ser65 upon OA-treatment, a PINK1-mediated post-translational modification essential for mitochondrial recruitment and Parkin activation^31^, showed a time dependent increase following OA treatment. To visualize PINK1 activation on depolarized mitochondria, we performed immunostaining on OA-treated WT and p.Gln336Arg fibroblasts for Ub^p-S65^, TOM20 and LAMP1 (Supplementary File 5F). Quantification of Ub^p-S65^ signal on mitochondria indicated robust PINK1 activation (Supplementary File 5G; Figure 5C). However, colocalization analysis of Ubp^S65^ with lysosomes revealed a significant decrease in cells carrying the EPG5 p.Gln336Arg variant (Supplementary File 5H), indicating defective delivery of phospho-ubiquitinated mitochondria to lysosomes.

We next investigated whether Parkin is recruited to damaged mitochondria during OA-induced mitophagy by co-transfecting with mt-Keima and GFP-Parkin. Delivery of mt-Keima to lysosomes at 24h-OA-treatment results in the formation of bright punctae with high ratio of excitation at 543/458 nm in response to the acidic microenvironment in the lysosome^32^. Initially, GFP-Parkin was localized diffusely throughout the control fibroblasts, while p.Gln336Arg fibroblasts showed significant increase in GFP punctae colocalizing with a low F_543_/F_458_ mito-Keima signal (Supplementary File 5I-K). GFP-Parkin recruitment was further increased in p.Gln336Arg cells after 24h OA-treatment (during late mitophagy, Figure 5E and Supplementary File 5L-M). A lower GFP-Parkin signal colocalizing with high F_543_/F_458_ mt-Keima signal in WT suggested that the fusion protein has been delivered to and degraded in mitolysosomes. To confirm the delivery of Parkin-positive mitochondria to lysosomes, cells were transfected with mCherry-Parkin followed by OA-treatment and immunostaining for mitochondrial matrix protein, citrate synthase (CS) and LAMP1 (Figure 5F). Quantification of Parkin-positive CS punctae colocalizing with lysosomes, identified as mitophagy events, were significantly decreased in p.Gln336Arg cells, indicating defective degradation and accumulation of Parkin-activated mitochondria (Supplementary File 5N).

We also examined PINK1/Parkin-dependent mitophagy in immortalized MEFs from *Epg5*^*Q331R*^ homozygous mice. We used transmission electron microscopy to identify the downstream block in mitophagy by analyzing mitophagosomal structures in MEFs stably expressing GFP-Parkin (Supplementary File 5O). Consistent with the increased accumulation of mitophagosomes observed in patient fibroblasts, *Epg5*^*Q331R*^ MEFs showed a significant increase in number of mitophagosomes during PINK1/Parkin mitophagy in the presence of BafA, compared to WT (Supplementary File 5P). Notably, mitophagosomes formed in *Epg5*^*Q331R*^ MEFs contained correctly sequestered mitochondria albeit those were significantly larger than in WT cell lines (Supplementary File 5Q), further supporting impaired lysosomal fusion of normally formed lysosomes. In MEFs stably expressing GFP-Parkin and mt-Keima, quantitative mitophagy measurements by FACS analysis showed a significant increase in mt-Keima spectral shift upon OA-induced mitophagy in WT, which was reversed by co-treatment with BafA (Supplementary File 5R). *Epg5*^*Q331R*^ homozygous MEFs also had a largely attenuated mt-Keima spectral shift in response to OA (Supplementary File 5S), again indicating lower levels of PINK1/Parkin-mitophagy.

Taken together, cellular data from fibroblasts derived from patients and mice with *EPG5* pathogenic variants indicate significant mitophagy and mitochondria alterations that may provide a cellular mechanistic explanation for the parkinsonism observed in *EPG5*-RDs, considering the crucial role of impaired mitophagy and mitochondrial quality control in genetic forms of PD.

## DISCUSSION

Here we present genetic, clinical and pathological data from the largest cohort of patients with pathogenic *EPG5* variants reported to date, complemented by cellular, *C. elegans*, and *M. musculus* models of EPG5 defects, replicating specific aspects of the human disease phenotype.

We provide evidence for an expanding range of phenotypes associated with EPG5 defects, suggesting a relatively milder end of the clinical spectrum different from classic VS^7^. Based on these observations, we propose to reserve the term “Vici syndrome (VS)” only for patients who fulfil at least 5 out of the 7 previously defined diagnostic criteria – callosal agenesis, (acquired) microcephaly, cataracts, cardiomyopathy, (combined) immunodeficiency, hypopigmentation and failure to thrive^11^ – and to use the term “*EPG5*-related disorder (*EPG5*-RD)” for all other patients with pathogenic *EPG5* variants without classic VS^7^. Whilst classic VS may be rare, our findings suggest that other *EPG5*-RDs may be more common, including more frequent presentations such as non-specific neurodevelopmental delay, epilepsy, spasticity, dystonia, and early-onset parkinsonism. Despite distinct features at different ends of the *EPG5*-related spectrum, the clinical continuum is evidenced for example by the more subtle callosal abnormalities at the milder end and the presence of epilepsy, spasticity, and dystonia as additional features in patients with classic VS, often overshadowed by the more striking syndromic features.

Not unexpectedly, we found that patients with classic VS were typically found through *EPG5* Sanger sequencing to be in a homozygous or compound heterozygous allelic state for at least one truncating variant, resulting in reduced EPG5 protein expression, whereas those with less specific clinical diagnoses before genetic testing were typically identified through an unbiased exome/genome sequencing approach and more often found to be homozygous or compound heterozygous for missense variants, expected to result in dysfunctional EPG5 protein. These observations suggest a dosage effect in *EPG5*-RD, with the phenotype of classic VS associated with marked reduction of the EPG5 protein and relatively milder phenotypes associated with expression of a dysfunctional EPG5 protein. A dosage effect is also suggested by the previous preliminary observation of early-onset neurodegeneration in heterozygous *EPG5* variant carriers in a small number of families, and the suggestion of *EPG5* variants as a modifying factor in Alzheimer’s disease and ALS^11,33,34^. Interestingly, an *epg5*-related dosage effect was also seen in a *Drosophila* model where heterozygous knockout and knockdown *epg5* flies displayed a milder phenotype compared to homozygous knockouts^28^. It is currently unclear if the suspected dosage effect correlates directly to a corresponding impairment in autophagic clearance, or alternatively the disturbance additional EPG5 roles in intracellular trafficking processes. Interestingly, a similar constellation is seen with the phosphoinositide-5 phosphatase gene *FIG4* also involved in intracellular trafficking, where biallelic null variants cause the severe neurodevelopmental disorder Yunis-Varon syndrome, while compound heterozygosity for truncating and missense variants cause the milder childhood-onset Charcot-Marie-Tooth disease 4J, and heterozygous deleterious variants cause a rare form of late-onset ALS^35-37^.

Whilst pathogenicity ascertainment is relatively straightforward for genotypes involving unequivocally pathogenic *EPG5* truncating variants, genotypes involving one or two missense variants are more difficult to ascertain. To address these difficulties, we subjected all missense variants to a rigorous *in silico* bioinformatic assessment and modelled the structural effects of selected missense variants applying crystallographic and computational tools; although the precise EPG5 protein structure is currently only still imperfectly understood, such modelling predicted significant consequences in regions of putative structural importance (Supplementary File 1). Moreover, for selected variants such as the recurrent EPG5 p.Gln336Arg variant, cellular studies revealed specific impairments of autophagic and mitophagic clearance confirming pathogenicity (see below).

We identified specific novel clinical and radiological presentations within the spectrum of *EPG5*-RDs that mirrored a number of monogenic disorders with dystonia, hereditary spastic paraplegia, and early-onset parkinsonism. On the clinical level, the overlap between these disorders is indicated by the emerging prominence of movement disorders in *EPG5*-RDs, in particular dystonia, also prominent with WDR45 defects, and spasticity, the clinical hallmark of autophagy-related forms of Hereditary Spastic Paraplegia due to variants in *SPG11, SPG15* and *SPG49*.

The neuroradiological ‘eye of the tiger’ sign, both reported here for the first time in *EPG5*-RDs, have previously been reported as part of the MRI spectrum in *PKAN*-or *WDR45*-associated neurodegeneration with brain iron accumulation^38^. On the radiological level, brain iron accumulation is a hallmark not only of WDR45 defects but may also occur in the later stages of various forms of HSP^39^. Vice versa, early radiological features of *WDR45* variants may closely mimic those of VS, further supporting not only a clinical but also a radiological continuum between those disorders^40^. To follow-up on a possible neuropathological basis for these observations, we suggest postmortem tissues investigations to analyze for brain iron accumulation in individuals with *EPG5*-RD. Along similar lines, the Lynx sign also reported here for the first time in *EPG5*-RDs is also a recognized feature of *SPG11*-and *SPG15*-related disorders^41^. Lastly, evidence of progressive cerebral atrophy is common in a range of autophagopathies, for example in Kjellin syndrome, an extension of the SPG15 spectrum^42^.

Albeit novel, this observed overlap of EPG5 defects with disorders of iron is not unexpected, considering the crucial role of autophagy and vesicular trafficking in normal iron homeostasis^43,44^. Considering that the iron-dependent regulated cell death mechanism ferroptosis is blocked by glutathione peroxidase 4 (GPX4)^45^, any deficit in autophagy flux may conceivably cause iron accumulation due to dysregulated feedback mechanisms^43,46^.

As an extension of our previous work on *EPG5*-related classic VS^11^, this expanded *EPG5*-RD cohort suggested prominence of motor disorders with either i) already postnatal or infantile onset, or, more commonly, ii) adolescent-onset dystonia and parkinsonism on the background of non-specific neurodevelopmental delay, with evidence for an underlying mitochondrial cytopathy both on the histopathological and cellular level. The complementary EPG5-defective organisms studied here are established models for motor disorders, and their sequential use in the paradigmatic example of a monogenic autophagy disease provides a comprehensive perspective on perturbed autophagosome-lysosome fusion, mitochondrial pathology and/or the associated movement disorders^30,47^.

Compared to the previously published *Epg5* knockout mouse model, our *Epg5*^*Q331R*^ mouse has a milder phenotype with an age-dependent motor phenotype, corresponding to but not fully replicating the human phenotypes associated with *EPG5* missense variants^29^. Interestingly, the phenotype in the *Epg5*^*Q331R*^ mouse was more suggestive of a neurological motor impairment with additional behavioural changes suggestive of a neurodegenerative disorder rather than a primary neuromuscular presentation. For the first time, and in contrast to *Epg5* knockout mice^29^, we could demonstrate that the autophagy defect in our knock-in model affects different central nervous system regions selectively, in a pattern comparable to humans with *EPG5*-RDs. Other important differences are that weight loss, often observed with neurodegeneration, does not occur in our knock-in mouse model, while it is already present after a few months in the *Epg5* knockout mice^29^ and, interestingly, in humans at the severe end of the *EPG5*-RD spectrum.

Early-onset parkinsonism as part of *EPG5*-RDs corresponds to earlier preliminary observations of an apparently increased Parkinson’s disease (PD) risk in heterozygous *EPG5* variant carriers in a relatively small cohort of families, and similar observations in other lysosomal storage disorders such as Gaucher disease, where the heterozygous carrier state is also associated with an increased PD risk^48^. While some phenotypic aspects in *Epg5*^*Q331R*^ mice resemble phenotypes observed in mouse models of MPTP-induced parkinsonism (reduced stride length, imbalance, and increased levels of anxiety)^49-52^, we were unable to fully investigate any phenotype worsening because, according to UK regulations, these mice had to be euthanized at ∼12 months of age for their recurrent spontaneous seizures^28^.

However, a putative link between *EPG5* variants and parkinsonism is supported by our previous observation of a specific age-related loss of dopaminergic neurons in a *epg5* knockout *Drosophila melanogaster* model^28^, and of a hypokinetic locomotion disorder in a *C. elegans epg-5* knockdown model reported in the present study. In *C. elegans*, we also observed an increase in spare oxidative capacity with knockdown of genes involved in autophagosome-lysosome fusion (*epg-5/EPG5, rab-7/RAB7, ccz-1/CCZ1*), similar to what is observed with knockdown of the mitophagy gene *pdr-1/PRKN*, but not in control worms. Such elevated ‘hyperactive’ mitochondrial electron transport indicates an excess of uncoupled capacity, in line with previous reports in humans and worms showing that *pdr-1*/*PRKN* knockout is associated with an increased spare capacity and more fused mitochondrial networks at L4 larval worm stage^53^; lymphoblasts from patients with Parkinson’s disease have also previously been shown to have increased spare capacity independent of patient age^54^.

Moreover, *in vivo* flux measurements in *epg-5i* worms showed increased LGG-1/LC3:RFP autophagosome punctae (indicator for impaired autophagic clearance)^55^ and DCT-1:GFP mitophagosome punctae indicative of stalled selective mitochondrial clearance. As PDR-1/PRKN positively regulates mitophagy and interacts with key mitophagy protein DCT-1/BNIP3^26^, our experiments indicated impaired mitophagic clearance in knockdowns of both *pdr-1/PRKN* and *epg-5/EPG5*. As these genes are highly conserved throughout evolution, we argue that human EPG5 defects are also associated with impaired mitophagic clearance, ultimately resulting in neurodegeneration and perturbed locomotion.

Finally, and along similar lines, cellular studies in human fibroblasts carrying recurrent homozygous *EPG5* missense variants confirmed disturbances of PINK1/PRKN-mediated mitophagy, thus providing a potential mechanistic explanation for the link between *EPG5* and parkinsonism. These *EPG5*-related findings correspond to observations in *PARK2*-and *PINK1*-mutated patient cells and induced dopaminergic neurons, showing accumulation of damaged mitochondria upon induction of mitophagy^56^. Histochemical analysis of substantia nigra (SN) sections from *PARK2*-and *PINK1*-mutated patients also revealed pS65-Ub punctae colocalizing with mitochondria and lysosomes similar to what we observed in *EPG5*-mutated cells^57^. The impaired mitophagy observed in *EPG5*-related patient cells also closely mimics mitochondrial abnormalities seen in a mouse model of Gaucher disease, a lysosomal storage disorder associated with PD in heterozygous *GBA* variant carriers^58^.

Genetic defects in several other autophagy components have already been linked to neurodegenerative disorders, indicating that well-maintained autophagic flux associated with normal EPG5 function is crucial for the normal function and maintenance of the central nervous system^11,59-62^. A widely acknowledged hypothesis regarding the ‘atypical form’ of parkinsonism, an emerging important feature of these disorders, with its rapid progression and cognitive decline is that massive protein and organellar accumulation may lead to excessive neuronal waste and degeneration^63^. Previous reports identified autophagopathies such as WDR45 defects as the cause of both childhood NDD^46^ and early adulthood parkinsonism^64^, suggesting substantial clinico-pathological overlap reflective of the molecular interactions between EPG5 and WDR45 in the autophagosome-lysosome fusion machinery (Figure 6). The autophagosome-lysosome fusion machinery is rapidly emerging as a hotspot for neurological disease, with pathogenic variants also in *SNAP29, RAB7A*, and HOPS complex components implicated in a wide range of neurological diseases^65-68^, highlighting a promising target for future investigations.

**Figure 6.**
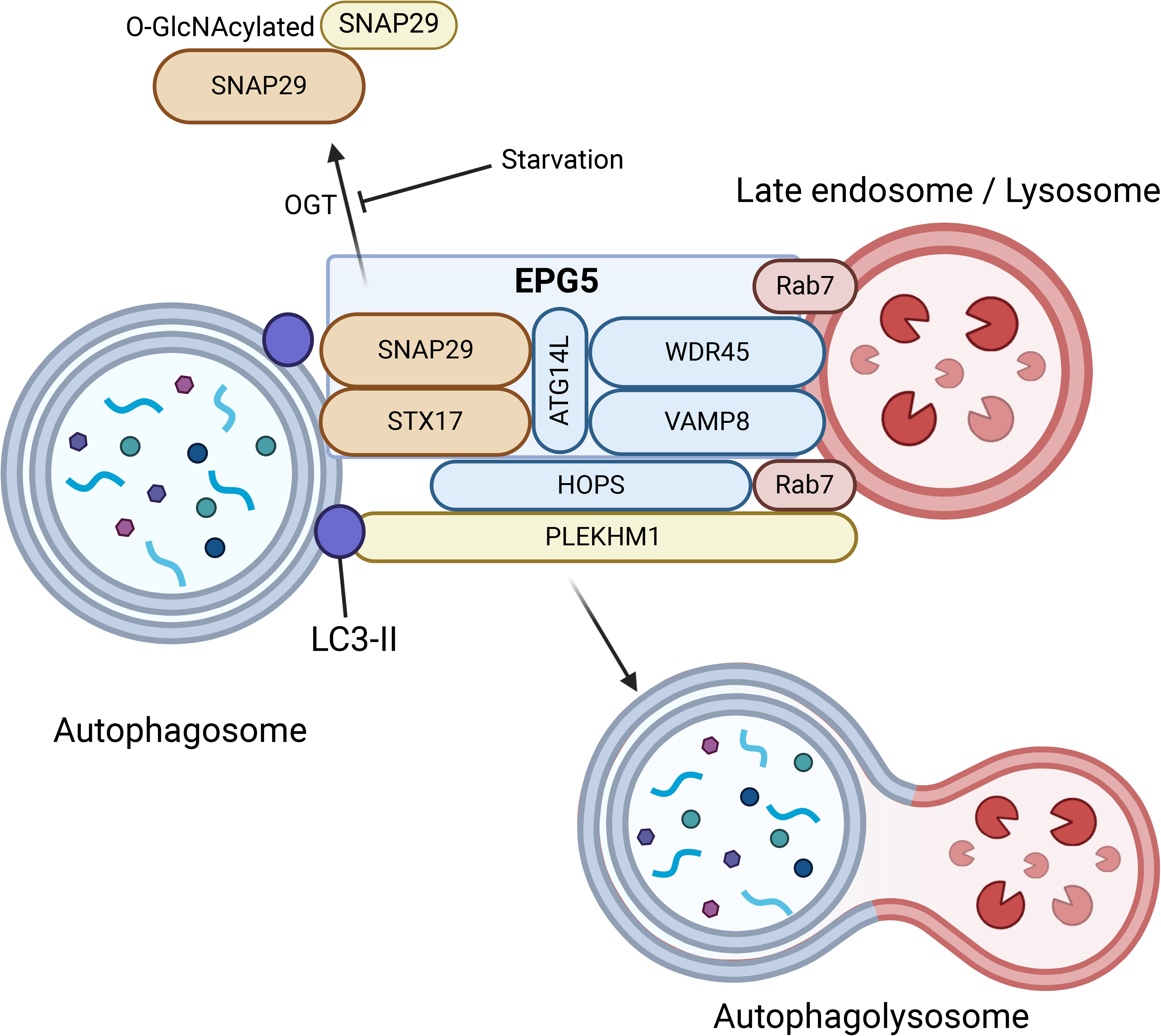
The autophagosome-lysosome fusion machinery as a hotspot for neurological disease. The fusion between autophagosomes and lysosomes in the final step of the autophagy pathway is a complex process that involves an intricate molecular machinery. In addition to EPG5, other proteins interacting within the autophagosome-lysosome fusion machinery have already been implicated in neurodevelopmental and neurological conditions or are candidates for as yet unresolved conditions. EPG5 = Ectopic P-Granules 5 Autophagy Tethering Factor; SNAP29 = Synaptosome Associated Protein 29; STX17 = Syntaxin 17; ATG14L = Autophagy Related 14; VAMP8 = Vesicle Associated Membrane Protein 8; Rab7 = Ras-related protein Rab-7a; HOPS = homotypic fusion and protein sorting-tethering complex; PLEKHM1 = Pleckstrin Homology And RUN Domain Containing M1; LC3-II = LC3-phosphatidylethanolamine conjugate.

## CONCLUSION

Here, we describe the largest cohort of *EPG5*-mutated patients reported to date, indicating a wide and expanding spectrum with novel phenotypes characterized by prominent movement disorders, in particular dystonia and early-onset parkinsonism. Our data from patient cells and various complementary model organisms mechanistically explain these findings, replicating various aspects of the human neurodegenerative and motor phenotypes, and disturbances of autophagic flux as well as mitochondrial function and mitophagy. This work will inform genetic counselling for patients and families affected by *EPG5*-RDs and may provide the basis for the development of targeted therapeutic options.

Our recent findings provide context to our previous preliminary observations of an increased PD frequency in (putative) heterozygous variant carriers in *EPG5* families, suggesting that the heterozygous allele carrier state may predispose to PD later in life; this hypothesis may warrant further investigation through massively parallel sequencing for variants in *EPG5* and other autophagy genes with higher minor allele frequencies in PD cohorts.

Taken together, these findings suggest a lifetime continuum of disease related to *EPG5* deficiency in humans, as well as an increasing overlap between various congenital disorders of autophagy.

## Data Availability

All data produced in the present study are available upon reasonable request to the authors.

## Abbreviations

EPG5: ectopic p granule protein 5
VS: Vici syndrome
MRI: magnet resonance imaging

## Acknowledgements

This work was supported by grants from the European Union Horizon 2020 Programme (765912 — DRIVE — H2020-MSCA-ITN-2017) to CD, MF and HJ, Action Medical Research (2446) to HJ and MF, and Action Medical Research (GN2959) to KS and MRD. HSD was supported by the Koeln Fortune Program/Faculty of Medicine, University of Cologne (371/2021 and 243/2022), as well as the Cologne Clinician Scientist Program/Medical Faculty/University of Cologne and German Research Foundation (CCSP, DFG project No. 413543196). AA was supported by the Max Planck Gesellschaft. TSB was supported by the Netherlands Organisation for Scientific Research (ZonMw Vidi, grant 09150172110002), and acknowledges ongoing support from EpilepsieNL and CURE Epilepsy. KÕ is supported by Estonian Research Council grant PRG2040. Funding bodies did not have any influence on study design, results, and data interpretation or final manuscript. Some of the authors of this publication are members of the European Reference Network on Rare Congenital Malformations and Rare Intellectual Disability ERN-ITHACA. [EU Framework Partnership Agreement ID: 3HP-HP-FPA ERN-01-2016/739516]. Biospecimens used in this article were obtained from the Northwestern Movement Disorders Center (MDC) Biorepository. As such, the investigators within the MDC Biorepository contributed to the design and implementation of the MDC Biorepository and/or provided data and collected biospecimens but did not participate in the analysis or writing of this report (Rizwan Akhtar, MD, PhD; Tanya Simuni, MD; Puneet Opal, MD, PhD; Steven Lubbe, PhD; Joanna Blackburn, MD; and Lisa Kinsley, MS, CGC). We like to thank the Coriell Institute for Medical Research (Camden, NJ, USA) and Dr Fleur Vansenne (Groningen, The Netherlands) for their kind gift of *EPG5*-mutated patient fibroblasts. We like to thank Dr Susan Byrne (Dublin, Republic of Ireland) for her past work on *EPG5*-related Vici syndrome.

## Appendix

### Figure and Table Legends

**Box 1. Clinical vignettes of illustrative patients from the milder end of the *EPG5*-related spectrum and with prominent movement disorder phenotypes, including early-onset parkinsonism. Age ranges neonate (0-1m), infant (1m-1y), toddler (1-3y), child (3-12y), adolescent (12-18y) and adult (> 18y)**.

A) This adolescent male compound heterozygous for *EPG5* variants p.Gly2231Val, and c.4474+1G>A presented in his adolescence with subacute onset of a complex movement disorder (including parkinsonism, generalized dystonia with orofacial dyskinesia and intermittent jerky action tremor) on the background of moderate neurodevelopmental delay; he progressed to almost complete tetraparesis within two years of disease onset. MRI findings included iron / mineral accumulation in pulvinar of the thalami (Figure 1Q).

B) This adolescent female compound heterozygous for *EPG5* variants p.Val464Ala and p.Tyr855Ter presented with severe progressive fluctuating generalized dystonia, parkinsonism and cognitive decline on the background of moderate neurodevelopmental delay. MRI of the brain demonstrated callosal agenesis, generalized atrophy, features suggestive of basal ganglia iron accumulation in substantia nigra and red nuclei (Figure 1N-O).

C) This adolescent female compound heterozygous for *EPG5* variants p.Arg1501Trp and p.Thr1994Ala presented with generalized dystonia and parkinsonism with cognitive decline followed by rapid deterioration and death at adolescence. She had a background of global developmental delay, congenital hydrocephalus and progressive optic atrophy. MRI revealed callosal dysgenesis, cerebellar atrophy, pontocerebellar hypoplasia, and thalamic involvement (Figure 1J-L).

D) This young adult female homozygous for the *EPG5* variant c.5943-9_5943-5del presented with parkinsonism with anarthria, an extraocular movement disorder, generalized dystonia pronounced in the oropharyngeal muscles, spasticity, an irregular jerky tremor in the lower limbs and cognitive decline. She had a background of global developmental delay, optic atrophy and complex partial seizures responsive to levetiracetam from toddler age. MRI performed at adolescence indicated generalized atrophy. She had a rapidly progressive course and died at adolescence.

E) These two siblings were homozygous for the *EPG5* variant p.Phe2004Ser and had a background of global severe neurodevelopmental delay. The older sibling developed dystonia, choreoathetosis, spasticity, dysarthria, parkinsonism with tremor, postural instability and hypomimia in adolescence, followed by rapid cognitive decline with sleep disturbance, anxiety, and death at young adulthood. MRI of the brain showed cerebral and cerebellar atrophy, callosal agenesis, Lynx sign, and pallidal T2-hyperintensities on MRI (Figure 1E). The younger sibling presented from adolescence with dystonia and parkinsonism followed by slow regression on a background of intellectual disability. She subsequently developed seizures responsive to phenytoin from young adulthood and died after further disease progression. MRI of the brain showed periventricular leukodystrophy.

F) These two sibling pairs who were first cousins from a large consanguineous family were homozygous for the *EPG5* p.Phe2287_Leu2288insPheProThrAlaGluPhe variant. All 4 patients presented with variable combinations of parkinsonism, dystonia, tremor and spasticity from their adolescent years on a background of global developmental delay and mild intellectual disability. MRI showed callosal dysgenesis, Lynx sign, generalized atrophy, and ventriculomegaly (Figure 1H). Course was progressive with cognitive decline in all, with one having died after further deterioration at young adulthood.

**Supplementary File 1**

**Additional methods and results**.

Supplementary methods include statistical analysis, protein modelling, and behavioural analyses of the *Epg5* Q331R knock-in mouse model. Methods for *Caenorhabditis elegans* assays include neuronal morphology analyses, locomotion analyses, and oxygen consumption assays. Methods for cellular assays include immunoflourescence, immunoblotting, generation of a GFP-Parkin and mt-Keima expressing stable cell line in MEFs, transmission electron microscopy (TEM), and mitophagy measurement using FACS. Supplementary results include data on protein modelling of EPG5 variants.

**Supplementary File 2**

**Proforma used for data capture**.

Blank clinical proforma with 207 items regarding phenotypic and genomic features of cohort patients. Y = yes, N = no, ND = not defined by collaborator, NA = information not available.

**Supplementary File 3**

**Summary of in silico assessments of the *EPG5* missense variants identified in our cohort**.

Ensembl Variant Effector Prediction analyses of 147 variants in *EPG5*: location according to GRCh38; consequence (splice, intron, missense, truncating variant); impact according to VEP (high, moderate, low); gene symbol (EPG5); exon number; intron number; HGVSc annotation; HGVSp annotation; cDNA position; CDS position; protein position; amino acid residue; codon annotation; SIFT rank score; PolyPhen rank score; CADD_PHRED score; BayesDel addAF rankscore; BayesDel noAF rankscore; ClinPred rank score; FATHMM converted rank score; GERP++ RS rank score; LRT converted rank score; M-CAP rank score; MPC rank score; MVP rank score; MetaLR rank score; MetaSVM rank score; MutPred rank score; MutationAssessor rank score; MutationTaster converted rank score; PROVEAN converted rank score; PrimateAI rank score; REVEL rank score; gMVP rank score; SpliceAI delta score acceptor gain; SpliceAI delta score acceptor loss; SpliceAI delta score donor gain; SpliceAI delta score donor loss; ADA score; RF score; gnomAD v3 genomes heterozygous allele count; gnomAD v3 genomes allele frequency; gnomAD v3 genomes allele number; gnomAD v3 genomes homozygous allele count; existing variation in dbSNP; ClinVar clinical significance annotation; ClinVar ID.

**Supplementary File 4**

(A) Above, chromatogram of an *Epg5* p.Gln331Arg/Q331R KI mouse sample. The dominant sequence is shown by the larger peaks and top sequence, which after the mutation corresponds to the beginning of intron 2. The sequence of the smaller peaks is shown by the bottom line, which after the mutation corresponds to the beginning of exon 3. Below, schematic presentation of the third, canonically spliced isoform of *Epg5*. The red arrows indicate the location of the Q331R mutation. (B) *Epg5* KI mice do not show changes in body weight at early stage (1.5 months) or endstage (∼12 months). (C) Muscle strength of mutant mice was not affected at endstage when measured in a grip strength assay, suggesting a neurological rather than a neuromuscular basis for the observed abnormalities. (D) All mice show reaching behaviour in the trunk curl test. (E) No head-bobbing was observed in any of the mice. (F) No difference in DPOAE threshold or (G) ABR threshold was detected between wildtype and *Epg5* KI mice. (H) Only 14.28% of the *Epg5* KI mice show a Preyer reflex in the acoustic startle test. (I) *Epg5* KI mice have a later turning response in the contact righting test. Data are presented as percentage in (D,E,H) and mean ± S.E.M. in (D,E,G). Data were analysed using multiple unpaired Student’s t-test. N=5-10 per group. ****p<0.0001.

**Supplementary File 5**

(A) and (B) Quantification of p62 and NDP52 levels from Figure 5A. Plots represents the data from three independent experiments. (C) Representative images of mRFP-GFP-LC3 transfected fibroblasts treated with rapamycin and/or bafilomycin for 24 h. Patient fibroblasts show increased accumulation of mRFP and GFP positive puncta (autophagosomes) in untreated and rapamycin condition. Scale bar: 10 μm. (D) Quantification of both RFP and GFP positive (yellow puncta, autophagososmes) and only RFP positive (red puncta, autolysosomes) LC3 puncta from the autophagy flux assay in C. Each experiment examined 20 transfected cells. (E) Immunostaining of healthy fibroblasts with outer mitochondrial marker Tom20, EPG5 and LAMP1 proteins, showing colocalization of EPG5 to fragmented mitochondria (colocalization mask) in response to oligomycin and antimycin-induced mitochondrial membrane depolarization. (F) To check the phopho-Ubquitylation (Ub^p-S65^) of mitochondria under OA condition, control and Q336R fibroblasts were immunostained for Ub^p-S65^, TOM20 and LAMP1. (G) and (H) Colocalization of TOM20, Ub^p-S65^and LAMP1 was analysed from n ≥ 20 immunostained cells. A significant decrease in colocalization events of LAMP1 and Ub^p-S65^ (indicated by arrowheads) is seen in Q336R fibroblasts with swollen mitochondria. Scale bars, 20 μm and 10 μm, inset, 5 μm. (I) Representative images of fibroblasts cells co-transfected with mito-Keima (ratiometric) and GFP-Parkin (gray scale) under untreated condition. (J) The proportion of the high ratio (561/458) signal area (red) to the total mitochondrial area plotted as mitophagy index. (K) and (L) Quantification of the number of GFP-positive puncta per cell. Each experiment examined n ≥ 20 transfected cells. Scale bar: 10 μm. (M) The Pearson coefficient indexes between GFP-Parkin and mito-Keima (high ratio area) shows the accumulation of GFP-Parkin on the mitophagosomes/mitolysosomes over time under normal and OA treated condition as shown in I) and Figure 5E. (N) Quantification of the number of mitophagy events in Figure 6F. (O) Representative TEM images of GFP-Parkin expressing WT and Q331R MEFs treated with OA and Baf A. Scale bar, 1 μm. (P) and (Q) Quantification of mitophagosomal number per square micron and area in WT and Q331R MEFs stably expressing GFP-Parkin. Plots represent the data from three independent experiments with n ≥ 20 TEM images. (R) FACS analysis of WT and Q331R MEFs stably expressing GFP-Parkin and mt-Keima. The P2 gated area encloses cells undergoing mitophagy and shows the percentage of cells within this gate of each plot. (S) The percentage of cells undergoing mitophagy. Plots represent the data from three independent experiments. Data represents the mean ± s.d. of at least three independent experiments and analysed by one-way ANOVA with Tukey’s multiple comparisons test. *P < 0.05, **P < 0.005, ***P < 0.001, ****P < 0.0001. ns: not significant.

## Notes

### Competing Interest Statement

The authors have declared no competing interest.

### Author Declarations

IRB of Medical Faculty, University of Cologne gave ethical approval for this work.

