## Supplementary File 1 for "Mutations in EPG5 are associated with a wide spectrum of neurodevelopmental and neurodegenerative disorders"

***EPG5* deficiency links neurodevelopmental and neurodegenerative movement disorders**

**Supplementary Material**

**Methods**

### Statistical analysis

All data were analysed using Prism 9 (GraphPad) Version 9.4.1 (458) and compiled as figures with Illustrator (Adobe). Unpaired Student’s t-test was used for comparison of 2 groups of normally distributed data. Analyses of variances (ANOVA) with Bonferroni’s multiple comparisons test was used for comparison of more than 2 groups.

**Molecular modelling of EPG5 missense variants**

A composite molecular structural model of human EPG5 was generated by determining a three-dimensional density map by single-particle cryo-EM and fitting predicted structural models of three EPG5 domains (hook, finger, shaft) predicted by AlphaFold2 ^1^ into this map. After manually reconnecting the domains, real space refinement was carried out using PHENIX ^2^ to correct for stereochemical errors of the full EPG5 structural model. To model different EPG5 disease variants, the mutagenesis function in the Pymol package version 2.5.5 (Schrodinger, LLC)  was used to identify a side chain rotamer that minimizes clashes with surrounding residues.

**Behavioural analyses of Epg5 Q331R knock-in mouse model.**

Staging of the mice was based on their performance in the rotarod assay, where mice were placed on a rotating rod for 5 minutes which accelerated from 4 to 40 rotations per minute and their latency to fall off the rod was measured. Rearing behaviour was observed and manually scored when the mice were allowed to roam around an open arena for 10 minutes. Virtually dividing the open arena in a centre zone and outer zone allowed us to measure their velocity in a specific zone of the arena. Muscle strength was measured by allowing the mice to grip onto a grip strength meter using front and hind paws together. The time it took to right themselves to an upward position was also measured when the mice were positioned on their back.

Auditory brainstem responses (ABRs) were measured using the method described in detail previously^3^. Briefly, mice were anaesthetised and placed in a sound-attenuating chamber (IAC Acoustics Limited) on a heating blanket. Subcutaneous recording needle electrodes (NeuroDart, Unimed Electrode Supplies Ltd) were inserted on the vertex of the head and behind both ears. Responses were evoked by calibrated broadband click stimuli (10 μs duration) and tone pips (5 ms duration, 1 ms onset and offset ramp) at increasing frequencies, at levels ranging from 0–95 dB SPL (in 5 dB steps) at a rate of 42.6 stimuli per second. Those responses (potentials) were then amplified, digitised, and bandpass-filtered between 300–3000 Hz, using custom software and hardware. Thresholds of ABRs were defined as the lowest stimulus level to evoke a visually identifiable waveform.

Distoriton Product Otoacoustic Emissions (DPOAE) were measured in the ear canal of the mice in response to sounds of two combined frequencies (f1 and f2) as described previously^3^. Measurements were made inside a sound-attenuating chamber (IAC Acoustics Limited) with the mouse positioned on a heating blanket. Mice were anaesthetised and were placed in a position where the head was tilted approximately 45 degrees so that the left ear was uppermost. A DPOAE probe containing a microphone and speaker drivers to generate tones was positioned vertically towards the opening of the ear canal. Continuous stimuli at increasing f1 and f2 frequencies were generated and presented using the probe. The f2 tone was presented at a frequency of 1.2 x f1, and a level 10 dB SPL lower than f1. Sound pressure levels of the f2 stimulus ranged from -10 dB to 65 dB in 5 dB steps. The amplitude of the DPOAE was then recorded during stimulus presentation and was digitised for analysis purposes. Several parameters were calculated such as the DPOAE amplitude, the mean noise-floor amplitude, and the standard deviation (SD) of the noise-floor mean. The threshold of the DPOAE was defined as the lowest stimulus level when the DPOAE amplitude exceeded the SD two times above the recording noise-floor.

To measure the acoustic startle reflex, a sound was presented to the mice and their response was observed which was defined by the presence or absence of a brief ear flick. The response was observed by a blinded observer (NI) and either noted as present or absent.

In the trunk curl test the mice are held up by their tail above a horizontal surface. Whether the mouse reaches out to the surface or whether it curls its trunk towards its tail is observed. The response was observed by a blinded observer (NI) and either noted as reaching or curling.

For the contact righting test single mice are placed in a Perspex tube (3 cm in diameter). This way the animal’s four feet are in contact with the bottom of the tube and its back is in contact with the upper side of the tube. The tube is then inverted so the animal is turned upside down. The time it takes the mouse to re-orientate itself in the tube is measured.

**Neuronal morphology analyses in *C. elegans***

We used the wdIs52 [F49H12.4::GFP + unc-119(+)] reporter strain for visualizing posterior lineage V somatosenory neurons. Worms were grown on RNAi bacteria from egg-on. Three independent biological replicates of 10 hand-picked larval stage 3 nematodes each were used for microscopy with 5μ L of 1M sodium azide. Images of nematodes with control RNAi (*luci*) and *epg-5i* were taken with Andor Dragonfly at 60x magnification at larval stage 3. We defined the primary, secondary, tertiary and quartenary branches of the PVD neuron according to previously published studies ^4,5^. We counted any abnormal branching within the first ten secondary branches per worm. Image titles were blinded to the analysing researcher.

**Locomotion analyses in *C. elegans***

RNAi (*luci* control, *epg-5i*, *rab-7i*, *ccz-1i*, and *pdr-1i*) was carried out from egg-on and worms were collected at day 1 adulthood. Swimming behavior and locomotion were analyzed in n = 10 worms for each of three biological replicates using the *C. elegans* swim tracking software CeleST**.**^6^ Stretch was measured as the maximal differences in curvature that occurred between the two most extreme curvature scores at any animal body part during a swimming stroke.^6^ Curling was measured as the relative percentage of time that an animal spent bent around its own body^6^.

### Oxygen consumption assay in a *C. elegans* model of *EPG5*-related disorders

**Oxygen consumption assay:** All experiments were performed with worms grown on RNAi bacteria for *luci* control, *epg-5i*, *rab-7i*, *ccz-1i*, and *pdr-1i* from egg-on to day 1 adulthood. Approximately 15 nematodes were picked by hand into each well of a 96-well Seahorse utility plate filled with M9 buffer. Three replicates from independent plates were used in one assay and the assay was performed twice, each on a different day. At least four wells per assay were used as blanks. For determining the maximum oxygen consumption rate and inhibiting mitochondrial respiration, FCCP and sodium azide were used at final concentrations of 15 µM and 10 mM, respectively. Basal and maximum oxygen consumption rate were measured using the Seahorse Respirometer (XFe96)45 at 25°C.

**Immunofluorescence**

Fibroblasts grown on coverslips, either untransfected or transfected with mCherry-Parkin (Addgene plasmid #59419) were treated with OA and fixed with 4% paraformaldehyde and permeabilised with 50 µg/ml digitonin in PBS for 10 min. Cells were then washed, blocked with 3% BSA and incubated with the following primary antibodies: EPG5 (ab122186), Citrate synthase (ab96600) Tom20 (sc-17764), LAMP1 (sc-20011), Phospho-Ubiquitin (Ser65) (#62802) in 3% BSA for 1h at RT followed by incubation with Alexa Fluor 488/594/633- conjugated secondary antibody for 1h at RT. Coverslips were mounted on glass slides and imaged as described in Materials and Methods. Mitophagy events/cell was quantified in Fiji by counting the mask of the colocalized regions from Parkin-CS mask and LAMP1 mask. 3D reconstruction was carried out using the Surface plugin of Imaris 8.4.2 (Bitplane). The Pearson coefficient index (R value) was quantified using Coloc 2 plugin in Fiji.

**Immunoblotting**

Fibroblasts were treated with DMSO, rapamycin (100 nM), bafilomycin (200 nM), oligomycin (2.0 µM) and antimycin A (2.0 µM) for the indicated time. Following treatment, cells were collected and homogenized in RIPA lysis buffer containing freshly added complete EDTA-Free protease (Roche) and Phosphatase Inhibitor Cocktail 2 (Sigma-Aldrich). All lysates were centrifuged and quantified using the Pierce BCA Assay Kit (Thermo Scientific #23227). Samples were separated on 12% Bis-Tris gels (#NW00122) immersed in MOPS running buffer (Invitrogen #NP0001) and transferred onto PVDF membranes (1620175, Bio-Rad) and subsequently probed with the following antibodies for protein detection: SQSTM1/p62 (Abcam, ab56416), CALCOCO2/NDP52 (Thermo Scientific, P422), LC3B ( Sigma, L7543), PINK1 (Abcam, ab23707), Phospho-Ubiquitin (Ser65) (Sigma-Aldrich, ABS1513-I), pan Ubiquitin (Santa Cruz Biotechnology, sc-166553), β-Actin (Cell Signaling,3700), HRP conjugated anti-rabbit and anti-mouse (Jackson ImmunoResearch, # 111-035-045, # 315-035-045). After incubation with secondary antibody, PVDF membranes were imaged by the BIO-RAD ChemiDoc XRS imaging system. Densitometry analysis was performed using Fiji.

**Generation of GFP-Parkin and mt-Keima expressing stable cell line in MEFs**

MEFs transfected with GFP-Parkin were cultured and selected in DMEM supplemented with G418 (500 μg/ml) until stable clones were visible. The stable clones were transferred to 96 well plate to obtain single clones using serial dilution method. These GFP-parkin single clones were used for TEM studies. To generate mt-Keima expressing MEFs, GFP-Parkin stable cells were transduced with pCHAC-mtmKeima-IRES-MCS2 (Addgene plasmid #72342) retrovirus for 24 h and selected for protein expression using fluorescence sorting.

**Transmission electron microscopy (TEM)**

GFP-Parkin MEFs cultured on coverslips and treated with OA to induce PINK/Parkin-dependent mitophagy. MEFs were co-treated with bafilomycin (Baf A) during OA-induced mitophagy to prevent mitophagosome turnover and thereby enable comparisons of mitophagosome number and size. After treatment, coverslips were fixed in EM fixative (2% glutaraldehyde + 2% paraformaldehyde in 0.1M sodium cacodylate) for 1 hr followed by washings with 0.1M Cacodylate Buffer. The Coverslips were then fixed in 1% osmium tetroxide and 1% potassium ferricyanide in 0.1M sodium cacodylate, followed by sequential dehydration in ethanol. The coverslips were embedded in Epoxy Resin (Araldite CY212) Kit (Agar Scientific Ltd.) according to the standard protocol. Embedded coverslips were sectioned to 50nm using an ultra-microtome fitted with a diamond knife, mounted onto TEM compatible copper grids. The grids were then stained with Lead Citrate for 3 min before proceeding for Imaging. Images were acquired using Jeol 1400 Transmission Electron Microscope. Images were taken at a magnification between 800X and 1200X (digital magnification).

Mitophagososmal structures were segmented using DeepMIB software^7^. To quantify mitophagosome area and number, images were imported in Fiji. Mitophagosome structures were manually traced to determine the area. To quantify mitophagosome numbers per square micron, a 16 × 16 μm grid was overlaid on each image to quantify mitophagosomal structures which included 90% of cytoplasmic area, and the numbers were presented per μm2 of cytoplasm.

**Mitophagy measurement using FACS**

MEFs stably expressing GFP-Parkin and mt-Keima treated as indicated and then resuspended in sorting buffer (145 mM NaCl, 5 mM KCl, 1.8 mM CaCl_2_, 0.8 Mm MgCl_2_, 10 mM HEPES, 10 mM glucose, 0.1% BSA). Measurements of mt-mKeima signal were made using BD LSRFortessa™ X-20 flow cytometer equipped with a 405-nm and 561-nm laser. Cells were excited with a violet laser (405 nm) with emission detected at 610 ± 10 nm with a BV605 detector and with a yellow-green laser (561 nm) with emission detected at 610 ± 10 nm by a PE-CF594 detector simultaneously. For each sample, 20,000 events were collected and gated for mt-mKeima positive cells using FACSDIVA software (v8.0.1). Data were analysed using FlowJo (v10, Tree Star).

**Results**

**Protein modeling**

For the p.Arg1293Cys mutation, the protein model shows that the sidechain of Arg1293 is buried within a hydrophobic core. This residue may be involved in hydrogen bond interactions with its neighboring residues. The substitution of Arg with Cys, which has a shorter sidechain, may disrupt these interactions and packing, destabilizing the local structure. For the p.Arg1785Cys mutation, the protein model shows that the sidechain of Arg1785 is buried within a hydrophobic core. This residue may be involved in hydrogen bond interactions with its neighboring residues. Replacing Arg with Cys may also disrupt these interactions and packing, leading to instability in the local structure. For the p.Arg1905Gln mutation, the protein model shows that Arg1905 is located at the tip of a helix adjacent to a flexible loop region. Potential hydrogen bond and hydrophobic interactions may be disrupted upon mutation into a shorter sidechain residue Glu. For the p.Cys2038Arg mutation, the protein model shows that Cys2038 is located in a loop region between two helices, potentially forming a disulfide bond interaction with Cys2084. Substituting Cys with Arg may disrupt the disulfide bond interaction and the positively charged sidechain may destabilize the helical bundle formation. For the p.Trp2097Cys mutation, the protein model shows that Trp2097 is located in a loop region between two helices and is buried within a hydrophobic core. Replacement of Cys may disrupt local interactions and packing in this region. For the p.Met2123Arg mutation, the protein model shows that Met2123 is located in the core of the helical bundle and surrounded by hydrophobic residues. The substitution of Met with Arg, which has a positive charge, may introduce charges that disrupt the local structure.

**Further Supplementary Files**

**Supplementary File 2**

**Proforma used for data capture.**

Clinical proforma with 207 items regarding phenotypic and genomic features of cohort patients. Y = yes, N = no, ND = not defined by collaborator, NA = information not available.

**Supplementary File 3**

**Summary of in silico assessments of the *EPG5* missense variants identified in our cohort.**

Ensembl Variant Effector Prediction analyses of 147 variants in *EPG5*: location according to GRCh38; consequence (splice, intron, missense, truncating variant); impact according to VEP (high, moderate, low); gene symbol (EPG5); exon number; intron number; HGVSc annotation; HGVSp	annotation; cDNA position; CDS position; protein position; amino acid residue; codon annotation; SIFT rank score; PolyPhen rank score; CADD_PHRED score; BayesDel addAF rankscore; BayesDel noAF rankscore; ClinPred rank score; FATHMM converted rank score; GERP++ RS rank score; LRT converted rank score; M-CAP rank score; MPC rank score; MVP rank score; MetaLR rank score; MetaSVM rank score; MutPred rank score; MutationAssessor rank score; MutationTaster converted rank score; PROVEAN converted rank score; PrimateAI rank score; REVEL rank score; gMVP rank score; SpliceAI delta score acceptor gain; SpliceAI delta score acceptor loss; SpliceAI delta score donor gain; SpliceAI delta score donor loss; ADA score; RF score; gnomAD v3 genomes heterozygous allele count; gnomAD v3 genomes allele frequency; gnomAD v3 genomes allele number; gnomAD v3 genomes homozygous allele count; existing variation in dbSNP; ClinVar clinical significance annotation; ClinVar ID.

**Supplementary File 4**

(A) Above, chromatogram of an Epg5 Gln331Arg/Q331R KI mouse sample. The dominant sequence is shown by the larger peaks and top sequence, which after the mutation corresponds to the beginning of intron 2. The sequence of the smaller peaks is shown by the bottom line, which after the mutation corresponds to the beginning of exon 3. Below, schematic presentation of the third, canonically spliced isoform of Epg5. The red arrows indicate the location of the Q331R mutation. (B) Epg5 Gln331Arg KI mice do not show changes in body weight at early (1.5 months) or end stages (12 months). (C) Muscle strength of mutant mice was not affected at end stage when measured in a grip strength assay, suggesting a neurological rather than a neuromuscular basis for the observed abnormalities. (D) All mice show reaching behaviour in the trunk curl test. (E) No head-bobbing was observed in any of the mice. (F) No difference in DPOEA threshold or (G) ABR threshold was detected between wildtype and Epg5 KI mice. (H) Only 14.28% of the Epg5 KI mice show a Preyer reflex in the acoustic startle test. (I) Epg5 KI mice have a later turning response in the contact righting test. Data are presented as percentage in (D,E,H) and mean ± S.E.M. in (D,E,G). Data were analysed using multiple unpaired Student’s t-test. N=5-10 per group. ****p<0.0001.

**Supplementary File 5**

(A) and (B) Quantification of p62 and NDP52 levels from Figure 6A. Plots represents the data from three independent experiments. (C) Representative images of mRFP-GFP-LC3 transfected fibroblasts treated with rapamycin and/or bafilomycin for 24 h. Patient fibroblasts shows increased accumulation of mRFP and GFP positive puncta (autophagosomes) in untreated and rapamycin condition. Scale bar: 10 µm. (D) Quantification of both RFP and GFP positive (yellow puncta, autophagososmes) and only RFP positive (red puncta, autolysosomes) LC3 puncta from the autophagy flux assay in C. Each experiment examined 20 transfected cells. (E) Immunostaining of healthy fibroblasts with outer mitochondrial marker Tom20, EPG5 and LAMP1 proteins, showing colocalization of EPG5 to fragmented mitochondria (colocalization mask) in response to oligomycin and antimycin-induced mitochondrial membrane depolarization. (F) To check the phopho-Ubquitylation (Ub^p-S65^) of mitochondria under OA condition, control and Q336R fibroblasts were immunostained for Ub^p-S65^, TOM20 and LAMP1. (G) and (H) Colocalization of TOM20, Ub^p-S65^and LAMP1 was analysed from n ≥ 20 immunostained cells. A significant decrease in colocalization events of LAMP1 and Ub^p-S65^ (indicated by arrowheads) is seen in Q336R fibroblasts with swollen mitochondria. Scale bars, 20 µm and 10 µm, inset, 5 µm. (I) Representative images of fibroblasts cells co-transfected with mito-Keima (ratiometric) and GFP-Parkin (gray scale) under untreated condition. (J) The proportion of the high ratio (561/458) signal area (red) to the total mitochondrial area plotted as mitophagy index. (K) and (L) Quantification of the number of GFP-positive puncta per cell. Each experiment examined n ≥ 20 transfected cells. Scale bar: 10 µm. (M) The Pearson coefficient indexes between GFP-Parkin and mito-Keima (high ratio area) shows the accumulation of GFP-Parkin on the mitophagosomes/mitolysosomes over time under normal and OA treated condition as shown in I) and Figure 6E. (N) Quantification of the number of mitophagy events in Figure 6F. (O) Representative TEM images of GFP-Parkin expressing WT and Q331R MEFs treated with OA and Baf A. Scale bar, 1 µm. (P) and (Q) Quantification of mitophagosomal number per square micron and area in WT and Q331R MEFs stably expressing GFP-Parkin. Plots represents the data from three independent experiments with n ≥ 20 TEM images. (R) FACS analysis of WT and Q331R MEFs stably expressing GFP-Parkin and mt-Keima. The P2 gated area encloses cells undergoing mitophagy and shows the percentage of cells within this gate of each plot. (S) The percentage of cells undergoing mitophagy. Plots represents the data from three independent experiments. Data represents the mean ± s.d. of at least three independent experiments and analysed by one-way ANOVA with Tukey’s multiple comparisons test. *P < 0.05, **P < 0.005, ***P < 0.001, ****P < 0.0001. ns: not significant.
