## Supplementary File 2 for "Mutations in EPG5 are associated with a wide spectrum of neurodevelopmental and neurodegenerative disorders"

| **General information** | FamNr |
| --- | --- |
|  | Patient Nr |
|  | Internal ID |
|  | Prev. Pub. Nr |
|  | *Surname* |
|  | *First name* |
|  | Sex |
|  | DoB |
|  | Current age or age at death |
|  | Alive or dead? |
|  | Diagnosis |
|  | Country |
|  | Contributor |
|  | Email |
| **Documentation** | Clinical images (Y/N) |
|  | Previous genetic testing (Y/N) |
|  | DNA available (Y/N) ? |
|  | Where ? (Details/NA/ND) |
|  | MRI available (Y/N) ? |
|  | Where ? (Details/NA/ND) |
|  | Fibroblasts available (Y/N/ND) |
|  | Where ? (Details/NA/ND) |
|  | Muscle biopsy available (Y/N) ? |
|  | Where ? (Details/NA/ND) |
|  | Other biosamples available (specify) ? |
| **References (if previously published) (Patient Nr in publication)** |  |
| **References (if previously published) (Patient Nr in publication)** |  |
| **References (if previously published) (Patient Nr in publication)** |  |
| **Family history (FH)** | Pedigree (Y/N) |
|  | Consanguinity (Y/N) |
|  | Affected relatives (Y/N) (Details) |
|  | FH Vitiligo (Y/N) (Details) |
|  | FH Cancer (Y/N) (Details) |
|  | FH Neurology (Y/N) (Details) |
|  | FH Twin pregnancies (Y/N/ND) (Details) |
|  | FH Other (Y/N) (Details) |
| **Genetics** | Testing (Type/Location) |
|  | Phase (homozygous/heterozygous) |
| ***EPG5* Variant 1** | Exon |
|  | DNA |
|  | Protein |
| ***EPG5* Variant 2** | Exon |
|  | DNA |
|  | Protein |
| ***EPG5* Variant 3** | Exon |
|  | DNA |
|  | Protein |
| **Presentation** | Presentation (Details/ND) |
| **Cause of death** | Cause of death (Details/NA/ND) |
| **Perinatal** | Foetal movements (Reduced/Increased/ND) |
|  | IUGR (Y/N/ND) |
|  | Gestation (Weeks) (ND) |
|  | Birth weight (kg) (Centile) |
|  | Birth length (cm) (ND) |
|  | Birth OFC (cm) (Centile) (ND) |
|  | Neonatal presentation (Detail) (ND) |
| **Development** | Motor development (Normal) (Delayed) (ND) |
|  | Sitting (age) (never) (ND) |
|  | Walking (age) (never) (ND) |
|  | Best motor ability (Detail) (ND) |
|  | Speech development (Normal) (Delayed) (ND) |
|  | Best language Skill (Detail) (ND) |
|  | Hearing (Normal/Abnormal/ND) |
|  | Hearing (Details) (ND) |
|  | Vision (Normal/Abnormal/ND) |
|  | Vision (Details/ND) |
|  | Cognitive Development (Normal) (Delayed) (IQ) (ND) |
| **Somatic development** | Current weight (kg) (ND) |
|  | Current weight (age) (Centile) (ND) |
|  | Failure to thrive (Y/N/ND) |
|  | FTT details (Details/NA/ND) |
|  | Current height (cm) (ND) |
|  | Current height (Age/Centile/ND) |
|  | Current OFC (cm) (ND) |
|  | Current OFC (Age/Centile/ND) |
| **General features** | Dysmorphic features (Y/N/ND) |
|  | Dysmorphic features (Details/ND) |
|  | Hypopigmentation (Y/N/ND) |
| **CNS involvement** | CNS involvement (Y/N/ND) |
|  | Brain MRI (Y/N/ND) |
|  | Brain MRI Age (ND) |
|  | MRI summary (Details/ND) (please attach separate report) |
|  | Callosal agenesis (Y/N/ND) |
|  | Cerebellar atrophy (Y/N/ND) |
|  | Pontocerebellar hypoplasia (Y/N/ND) |
|  | Thalamic involvement (Y/N/ND) |
|  | Heterotopia (Y/N/ND) |
|  | Schizencephaly (Y/N/ND) |
|  | Calcifications(Y/N/ND) |
|  | Iron deposition (Y/N/ND) |
|  | Microcephaly (Y/N/ND) |
|  | Other CNS abnormalities (Y/N/ Details/ND) |
|  | PM Brain (Y/N/ND) |
|  | PM Brain (Details/ND) |
| **Epilepsy** | Seizures (Y/N/ND) |
|  | Seizure onset (Age/ND) |
|  | Seizure Type(s) (Details/ND) |
|  | EEG Findings (Details/ND) (please attach formal report) |
|  | AED (Details/ND) Indicate if effective (E) or ineffective (I) |
|  | Ketogenic diet (Y/N/ND) Indicate if effective (E) or ineffective (I) |
|  | Status epilepticus (Y/N/ND) |
|  | Non-convulsive status (Y/N/ND) |
| **Movement disorder** | Movement disorder (Y/N/ND) |
|  | Movement disorder details (Details/NA/ND) |
|  | Dystonia (Y/N/ND) |
|  | Dystonia Onset (Age/NA/ND) |
|  | Myoclonus (Y/N/ND) |
|  | Myoclonus Onset (Age/NA/ND) |
|  | Spasticity (Y/N/ND) |
|  | Spasticity Onset (Age/NA/ND) |
|  | Parkisonism (Y/N/ND) |
|  | Parkinsonism Onset (Age/NA/ND) |
|  | MD Medication (Details/NA/ND) Indicate if effective (E) or ineffective (I) |
|  | CSF Neurotransmiitter (Normal/Abnormal/ND) |
|  | Other neurological features (Details/N/ND) |
| **Learning, behaviour and psychosocial features** | Cognitive abilities (Details/ND) |
|  | Psychiatric/behavioural abnormality (Details/N/ND) |
| **Muscle involvement** | Muscle weakness (Y/N/ND) |
|  | Muscle weakness details (Facial/Proximal/distal/axial/general/ND) |
|  | CK levels (IU/l/ND) |
|  | Muscle Biopsy (Y/N/ND) |
|  | Muscle biopsy Age (Age/ND) |
|  | Muscle Biopsy Details (Details/ND) Please attach report |
|  | Type 1 predominance (Y/N/ND) |
|  | Fibre Type disproportion (Y/N/ND) |
|  | Vacuoles (Y/N/ND) |
|  | Cores (Y/N/ND) |
|  | Respiratory chain enzyme (RCE) studies (Y/N/ND) |
|  | RCE details (Details/ND) |
|  | Electron microscopy (EM) (Y/N/ND) |
|  | EM Details (Details/ND) |
| **Nerve involvement** | Neuropathy (Y/N/ND) |
|  | DTR (Absent/Reduced/Increased/ND) |
|  | EMG (Normal/Abnormal/ND) |
|  | EMG Details (Details/ND) |
|  | Nerve biopsy (Y/N/ND) |
|  | Nerve biopsy Details (Details/NA/ND) |
| **Ocular involvement** | Ocular involvement (Y/N/ND) |
|  | Cataracts (Y/N/ND) |
|  | Cataracts Onset (Age/ND) |
|  | Optic atrophy (Y/N/ND) |
|  | Other ocular abnormalities (Details/N/ND) |
|  | VER (Y/N/ND) |
|  | VER Detail (Detail/NA/ND) |
| **Hearing involvement** | Hearing involvement (Y/N/ND) |
|  | Sensorineural deafness (Y/N/ND) |
|  | BAER (Y/N/ND) |
|  | BAER Details (Details/NA//ND) |
| **Cardiac involvement** | Cardiac involvement (Y/N/ND) |
|  | Cardiac involvement Detail (Detail/NA/ND) |
|  | Congenital heart defect (Y/N/ND) |
|  | CHD Type (Type/NA/ND) |
|  | Cardiomyopathy (CM) (Y/N/ND) |
|  | Onset CM (Age/NA/ND) |
|  | Type CM (Type/NA/ND) |
|  | Cardiac US (Y/N/ND) |
|  | Cardiac US Details (Details/ND) |
|  | Cardiac MRI (Y/N/ND) |
|  | ECG (Y/N/ND) |
|  | ECG Details (Details/ND) |
| **Pulmonary Involvement** | Pulmonary involvement (Y/N/ND) |
|  | Pulmonary involvement details (Details/NA/ND) |
|  | Interstitial lung disease (Y/N/ND) |
| **Thyroid involvement (Y/N/ND)** | Thyroid involvement (Y/N/ND) |
|  | Thyroid involvement details (Details/NA/ND) |
| **Thymus involvement (Y/N/ND)** | Thymus involvement (Y/N/ND) |
|  | Thymus involvement details (Details/NA/ND) |
| **Hepatic involvement** | Hepatic involvement (Y/N/ND) |
|  | Hepatic involvement details (Details/NA/ND) |
|  | Hepatomegaly (Y/N/ND) |
| **Renal involvement** | Renal involvement (Y/N/ND) |
|  | Renal involvement details (Details/NA/ND) |
|  | Renal tubular acidosis (Y/N/ND) |
|  | Electrolyte disturbance (Y/N/ND) |
|  | Electrolyte disturbance details (Details/NA/ND) |
| **Gastric involvement** | Gastric involvement (Y/N/ND) |
|  | Feeding difficulties (Y/N/ND) |
|  | Gastrostomy (Y/N/ND) |
|  | Enterocolitis (Y/N/ND) |
| **Immune system** | Immune system involvement (Y/N/ND) |
|  | Immune system involvement details (Details/NA/ND) |
|  | Immunodeficiency (Y/N/ND) |
|  | Immunodeficiency Type (B-cell/T-cell/Combined/ND) |
|  | Age at diagnosis (Age/NA/ND) |
|  | Number of febrile infections within 1st year of life (n/ND) |
|  | Serositits (pleuritis / peritonitis) (Y/N/ND) |
|  | Arthritis/uveitis (Y/N/ND) |
|  | Meningitis/ encephalitis (Y/N/ND) |
|  | Lymphadenopathy (Y/N/ND) |
|  | Allergies/eczema (Y/N/ND) |
|  | Measles-like rash (Y/N/ND) |
|  | Ectodermal dysplasia (Y/N/ND) |
|  | Ichthyosis (Y/N/ND) |
|  | Ear infections (Y/N/ND) |
|  | Skin infections (Y/N/ND) |
|  | Other infections (Details/NA/ND) |
|  | Amyloidosis, hyper-IgE (Y/N/ND) |
|  | Immunoglobulin results (Details/ND) |
|  | Inflammatory markers (Details/ND) |
| **Haemotology** | Haematological abnormalities (Y/N/ND) |
|  | Haematological abnormalities details (Details/NA/ND) |
|  | Anaemia (Y/N/ND) |
|  | Neutropenia (Y/N/ND) |
|  | Lymphopenia (Y/N/ND) |
|  | Thrombocytopenia (Y/N/ND) |
|  | Other haematological abnormalities (Details/NA/ND) |
| **Therapies trialled** | Therapies trialled (Details/NA/ND) |
| **Additional information** | Additional information 1 |
|  | Additional information 2 |
| **Date entry** | Date entry (Date/Initial) |
