## Supplementary figures and images for "Mutations in EPG5 are associated with a wide spectrum of neurodevelopmental and neurodegenerative disorders"

### Supplementary File 4

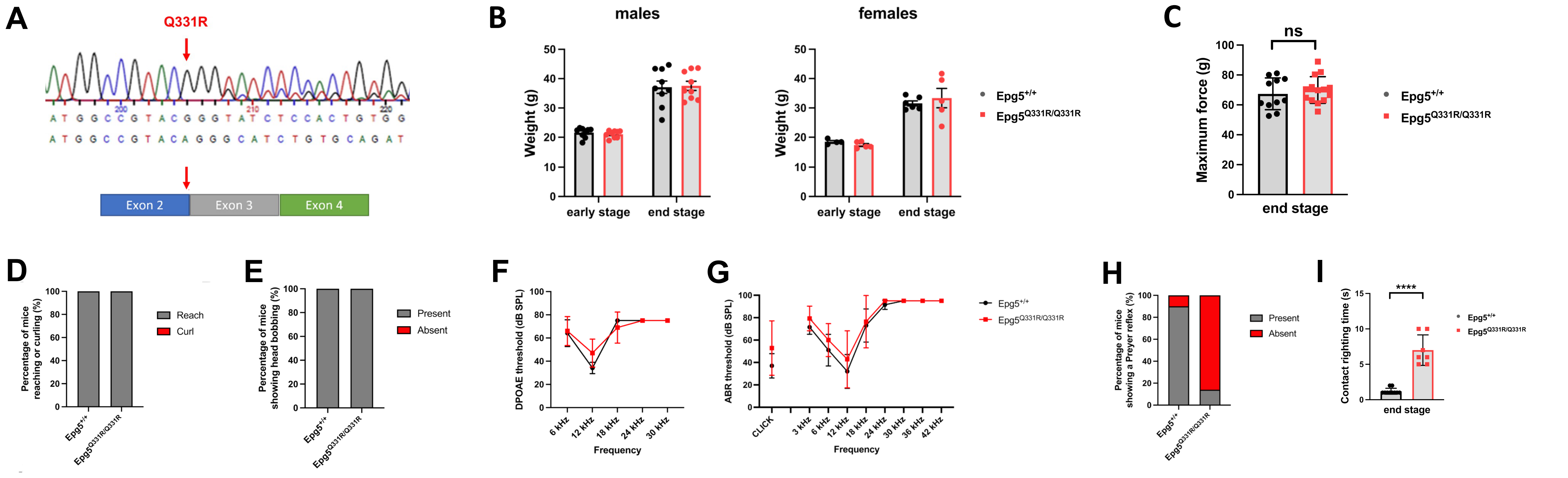

### Supplementary File 5

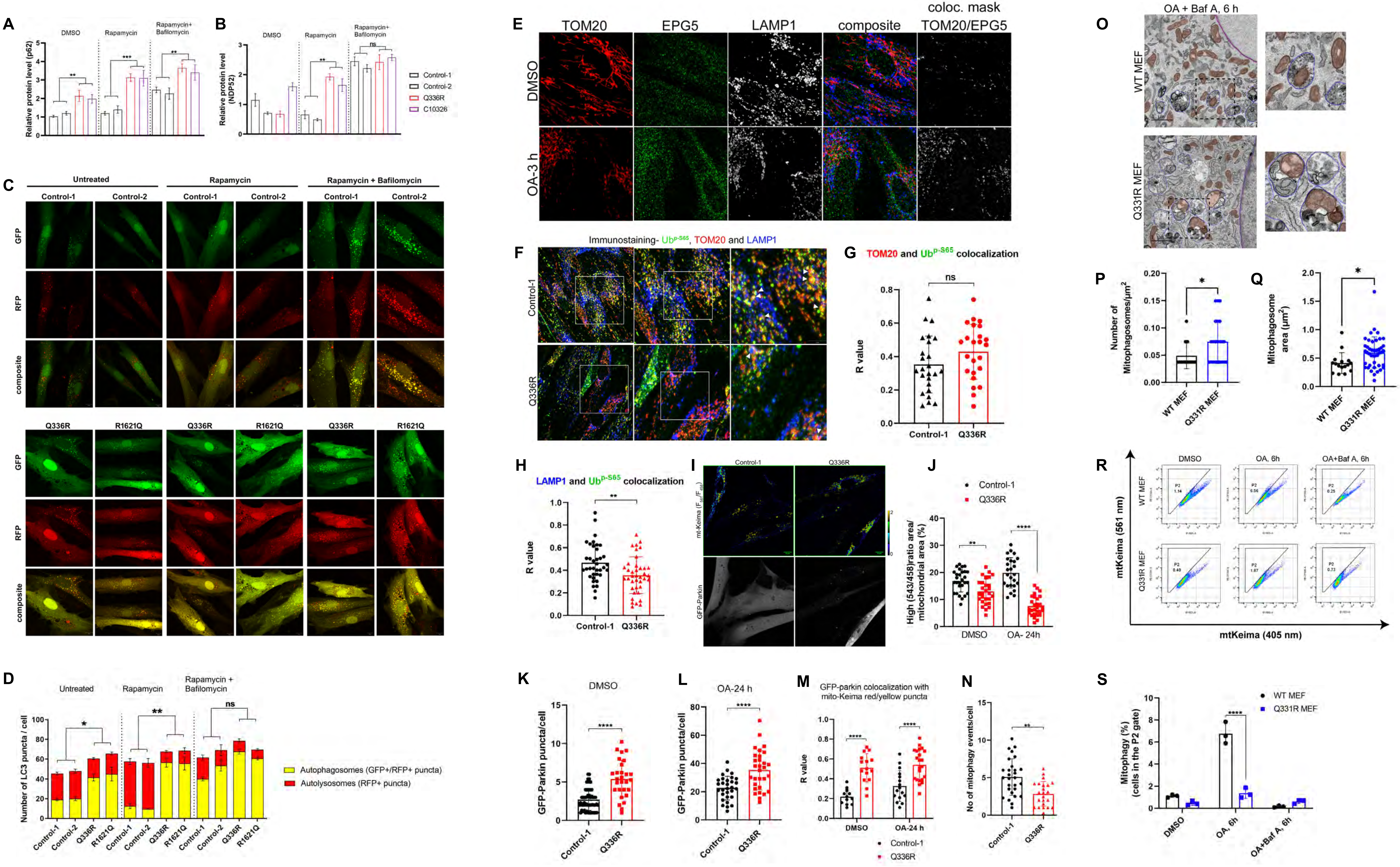
